# A network-based atlas of human skeletal muscle aging

**DOI:** 10.64898/2026.02.15.26346348

**Authors:** Tanner Stokes, Changhyun Lim, Muhammad Ali, Jonathan C. Mcleod, Jack Gisby, Katia Mariniello, Hannah Crossland, Jalil-Ahmad Sharif, Colleen Deane, Thomas C Moseley, Nasim M Ismail, ME Lixandrao, CH Volmar, Peter J McCormick, Robert J Brogan, James Whiteford, H Roschel, Stuart M. Phillips, Iain J Gallagher, Gregory Slabaugh, Bethan E Phillips, William E Kraus, Philip J Atherton, J Paul Chapple, James A. Timmons

## Abstract

Skeletal muscle metabolic and physical capacities are influenced by both genetics and load status and decline with age. Recent advances in sequencing have detailed cell types at unprecedented detail; yet these approaches do not scale to adequately model human muscle physiological heterogeneity. We produced a powerful resource for ageing studies, including consistent deep transcriptomic profiles of 1,675 human muscle biopsies (∼28,000 genes per profile) and multiple single-cell spatial transcriptomic technologies. We present several novel models of tissue ageing. Five Quantitative network models (QNMs), built using >40 trillion calculations and 930 human muscle transcriptomes, modelled aging and the influence of load status. Additional differential expression (DE) signatures for atrophy, hypertrophy and cardio-respiratory adaptation were integrated with single-cell RNAseq and cell-specific bulk profiles to reveal cell-enriched modules and the topology of human skeletal aging. Rapamycin transcriptomes from cultured muscle and endothelial cells, along with in vivo signatures for insulin resistance and sex, were integrated into these analyses. We show that >3,000 genes are DE with muscle age (equally up and down); that a novel pre-frailty signature in elderly subjects has a remarkably strong overlap with the response of healthy muscle during experimental atrophy and that the hypertrophy signature in elderly muscle, but not young muscle, opposes the age-regulated transcriptome. We report that non-responders for hypertrophy or gains in cardio-respiratory capacity have highly distinct genome-level response to exercise. QNM revealed cell-specific processes in endothelial cells and fibroblasts, including novel interactions between insulin sensitivity, age and senescence. From two hundred and eighty-six hub genes consistent in both young and old muscle network models, 27% had known roles in muscle biology, while of the top 50 hub genes (45% protein coding), 80% were newly linked to human muscle biology, including ARHGAP4, CEP131 and IFITM10 and many short- and long-noncoding RNAs. Many genes demonstrated extreme changes in topology in old muscle, such as the neddylation and aging linked gene, DCUN1D5. GeoMX-based spatial muscle fibre-type profiling (57 regions), along with Xenium (8 regions) and Merscope (54 regions) single-cell spatial technologies located key aging, frailty and load-responsive genes to individual cell types and provided novel insight into the location of autocrine/paracrine secreted factors such as GDNF, while IL6 was located to rare endothelial cells. A machine-learning model ranked the factors most associated with the topological changes with age. This prioritised network features over DE signatures, highlighting positive correlating edges to down-regulated genes during atrophy, genes up-regulated by Rapamycin and both positive and negative correlating insulin sensitivity features, along with gene hub status, best explained muscle ageing. Genome level modelling produced an independently validated transcriptomic ‘age clock’ and found it to be invariant to muscle load status in people >50y, while we revealed novel interactions between gene length and age. Release of an unprecedented level of consistently aligned genomic data, along with QNMs with >7,000 searchable modules, provides a powerful resource for the aging research communities.

## Introduction

Human resilience is underpinned by maintenance of musculoskeletal integrity. Human skeletal muscle phenotype is influenced by genetics^1,2^ and load status^3–5^ e.g. exercise activities. It is particularly challenging to apply genome-wide gene-expression technologies to human muscle^6^, partly because RNA responses are extremely divergent among individuals that demonstrate variable functional adaptability to altered load^2,7–9^. For cardiometabolic phenotypes, hundreds of individuals are required to accurately model the molecular processes shaping human muscle ^10–12^. These observations have led to significant efforts to pool gene-expression data from a wide variety of studies and genome-wide technologies^13,14^. Relying on two thousand diverse profiles, Myominer^13^ utilised accurate genomic realignment and consistent data quality control processes to provide the first robust large-scale meta-analysis of the human muscle bulk transcriptome but lacks coverage of the non-coding transcriptome, an influential component of network architecture^15^. Furthermore, while existing resources provide a convenient way to explore muscle gene expression, they lack quantitative network modelling^16^ and so do not reveal detailed network topology (e.g., how genes connect to each other and organize into higher-level modular structures). More recently, tissue atlas projects are revolutionising our understanding of tissue composition^17^. Recently, pioneering single-cell or nuclei RNA sequencing (scRNAseq and snRNAseq) has been used to characterise the single-cell contributions to human skeletal muscle tissue^18^. Intercostal muscle from seventeen adults was used for a pilot analysis of aging, to study the role of senescence, inflammation and metabolic dysfunction in muscle, finding most genes were down-regulated with age. This study^18^ also observed that <5% of these genes were also regulated in aged mouse skeletal muscle, emphasising the importance of human studies. Intercostal muscle is specialised and it’s unclear how molecular aging was influenced by the relatively constant loading. Further, myosin complex proteins were used^18^ to link nuclear transcriptomes with the major muscle fibre types (used to approximate for physiological function^19^) despite their probable lack of specificity for this task^20^.

Using anatomically diverse samples, Lai *et al* examined age-related changes in thirty-one people^21^ aged before or after the midlife age transcriptomic shift^11^. A subset of biopsies were subject to scRNAseq and snRNAseq, estimating cell composition from ∼80K cells, and reporting 1,873 genes per cell. This sparse coverage of the cell transcriptome limits the use of quantitative co-expression networks^22^. In contrast, the more complete bulk transcriptomic networks have correlation driven through cell-type expression patterns^23^. To date, no human muscle resource has taken advantage of these recent technological advances combined with the strengths of optimised bulk profiling^6^ to create a machine learning model of human skeletal muscle age interpreted by load-status^24^, longevity mechanisms^25,26^ and metabolic health. To address these gaps, we generated coding and noncoding transcriptomes from >1,500 muscle biopsies (**Figure 1**) obtained from people recruited in our clinical laboratories and developed a multi-omic machine learning driven model of human skeletal muscle aging. Critically, to avoid the limitations of cross-platform data integration, we utilised a single customised technology and pipeline to produce a signal optimised, accurately aligned, deep transcriptome. We illustrate the utility of these new human muscle models, by building a machine-learning based model of the network re-wiring of muscle with age, and identify novel relationships between gene length^27,28^, aging^27,29,30^ and protein coding status. We also present a new transcriptomic age “clock” and its response to altered load status in >500 people. Release of the raw data, and network models represents a powerful new resource to facilitate analysis of human skeletal muscle biology across age and sex.

**Figure 1.**
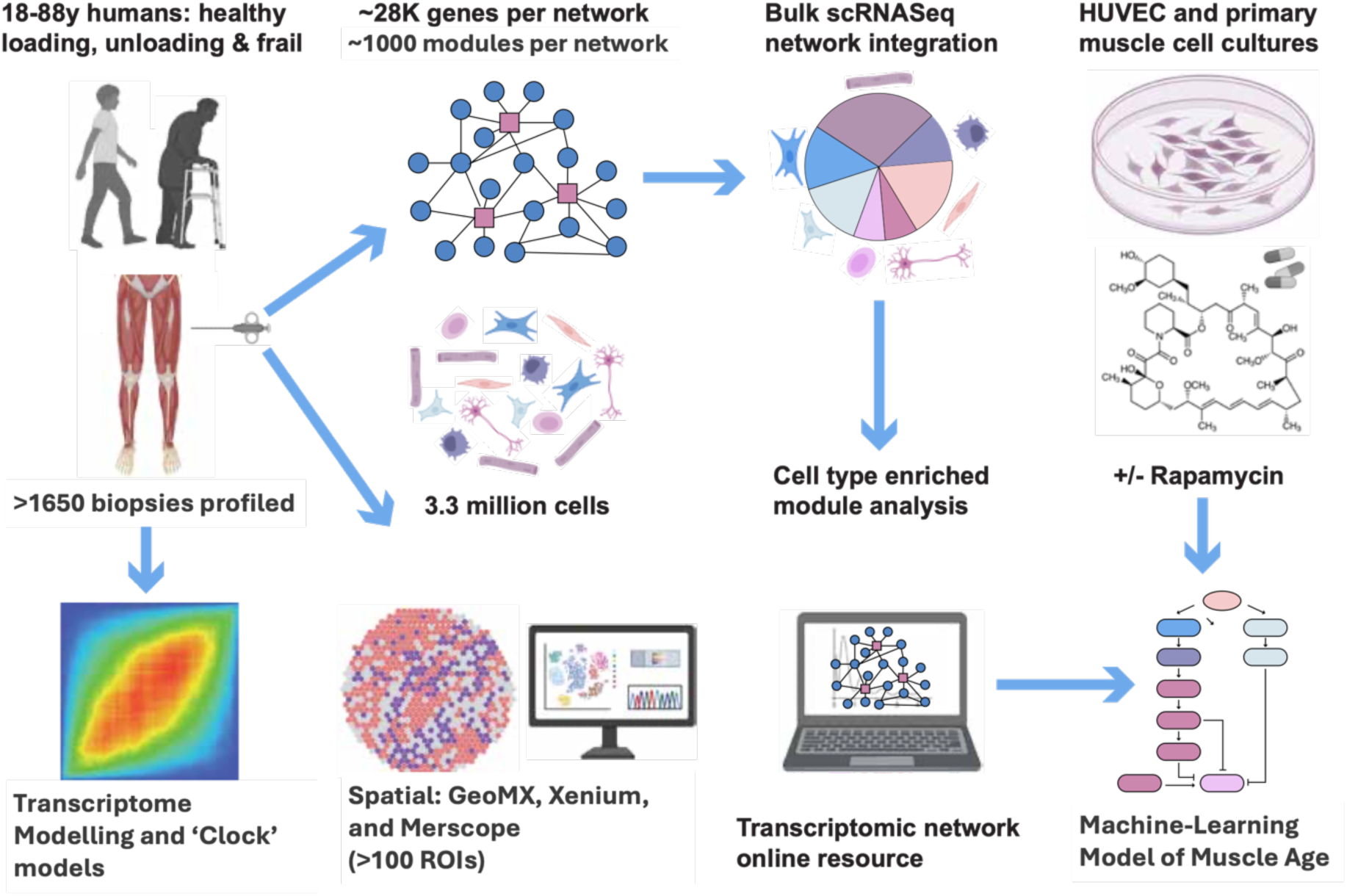
Project over-view diagram. We generated >1500 bulk transcriptomic profiles from biopsy samples obtained physiologically characterised humans, using a single platform and optimised methods^6,12,24,31–33^. This methodology captures a far more diverse coding and noncoding transcriptome from skeletal muscle than short-read RNA-sequencing^6,24,31^. Aligned to the current genome and transcriptome (and updatable against new genome releases), each transcript signal was optimised at the probe level (Figure S1A)^6,32^. The resource includes profiles of muscle from 18y to 88y, as well as muscle responding to hypertrophy, cardiorespiratory adaptation, short and long-term unloading induced atrophy, and 70y+ individuals with and without evidence of emerging frailty. We integrated scRNAseq, single cell type culture and bulk transcriptomics to identify cell type enriched QNM modules. Machine learning and regression-based models were used to develop tools to explain topological changes in muscle network structure with age and study the impact on load on tissue age predictions.

## Results and Discussion

We present a robust set of quantitative network models of human skeletal muscle across load and age, that reveal molecular events hidden in differential expression and pathway analysis. This new resource enables comparison of *consistently* measured RNA across distinct groupings of age, load and sex. We provide an unprecedented release of both raw data (>1,600 profiles) and computationally demanding network models, including >7,000 searchable network modules with single cell type enrichment profiles (Figure 2A-D, html link).

**Figure 2.**
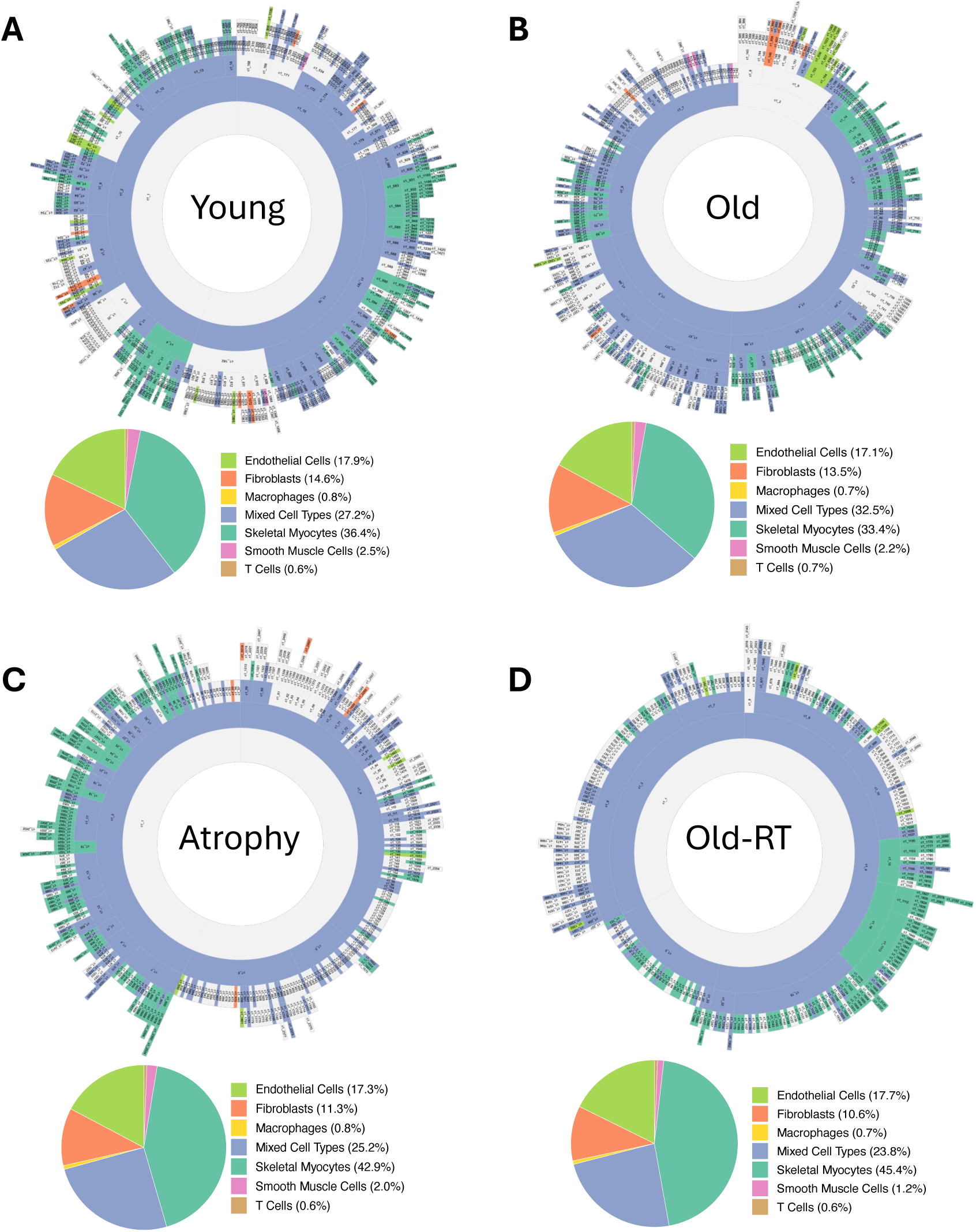
Hierarchal network structures and dominant cell types per module of human skeletal muscle. Presentation of a novel sunburst-based representation of the hierarchal organization of a MEGENA network. Each module is colored by its dominant cell type signature with ∼40% of the distal smaller modules being dominated as expected by myocyte gene expression, with the proximal large modules are mixed-cell type profiles. A) N=270 younger people (age=25.7), B) N=270 younger people (age=68.4y), 60 older people who were responders to resistance training (hypertrophy phenotype) and 47 adults subject to 1-2 weeks of knee-brace based immobilisation and atrophy of the quadriceps muscle.

### Implementing robust network modelling

Application of network modelling to large scale transcriptomic data reveals the underlying molecular and cell interactome, providing a method distinct from DE analysis, to study physiological regulation or aging mechanisms. QNM captures both the underlying cellular composition and transcriptional expression patterns of tissue-resident cell populations,^34,35^. It can infer the function of poorly-characterized genes via ‘guilt by association^34,35^. Furthermore, whereas scRNA-(and bulk) sequencing data is susceptible to stochastic and competitive mRNA capture, with substantial gene ‘drop out’, QNMs built with high sensitivity bulk transcriptomics captures lowly-expressed functionally relevant genes^6,31,34^. To build each QNM, we first implemented bespoke informatics reprocessing ^6,12,32^ of the high-sensitivity^36^ exon-level HTA bulk transcriptomic platform. Each of the 7M probes were aligned to the current genome and transcriptome (see methods) with ∼500,000 ‘dead’ probes removed from the ∼220,000 probe-sets (**Figure S1A**).

We utilised a subset of 952 bulk transcriptomic profiles to build 5 QNMs, representing transcriptomic profiles from ∼3.3 million cells (**Table 1**). The average coverage per cell from muscle single cell atlas analysis is ∼1,900 genes per cell, while our networks are built from >28,000 genes.

**Table 1.**
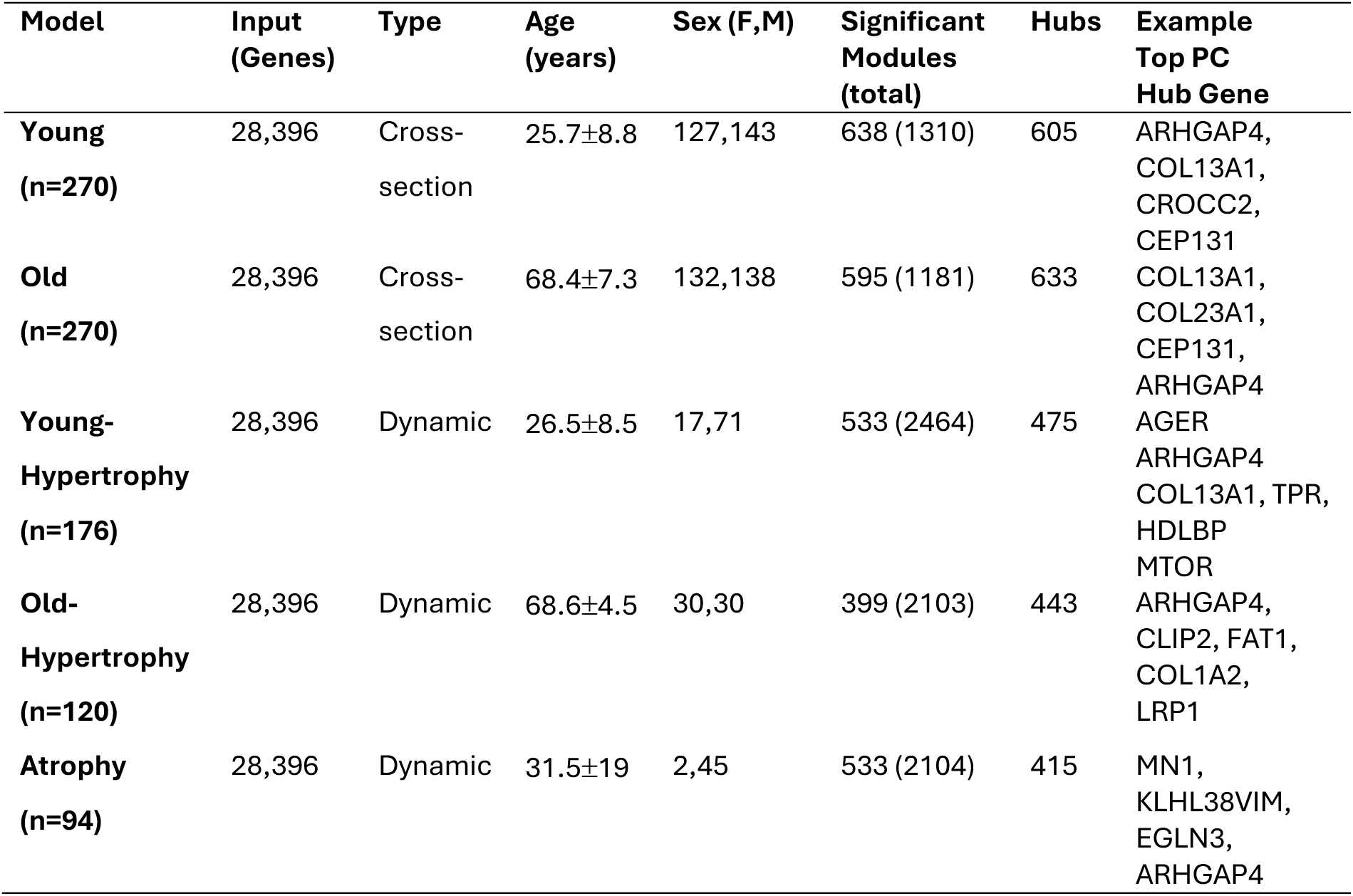
Input data demographics and network characteristics of the five Megena quantitative network models. The sample size is the number of biopsy profiles used for each network. Input is the number of rows in the QNM, where each gene is represented with the top graded transcript. The type of network refers to whether the model is built using cross-sectional data (e.g. 270 ‘young’ individuals below the age of 50y) or delta-gene expression values from a paired pre-post transcriptome.

Several algorithms exist to construct co-expression networks from transcriptomic data; to date however, most studies on human tissue use Weighted Gene Co-expression Network Analysis (WGCNA)^37^. WGCNA was developed at a time when computational resources were very limited, and it returns results for almost any sample size^38^ within a few minutes. Originally developed to model *differential* gene-expression, the WGCNA method is based on attempting to achieve a network with scale free properties^37,39^ and it has no robust statistical thresholding. Module membership can be indistinct from random^40^ and more importantly no module architecture is revealed. As it is known that strong to weak paired correlations require a sample size from ∼30 to 150 for reliable estimation^41,42^, and larger numbers of samples are required to generate a reproducible gene expression network,^43^ we implemented Multiscale Embedded Gene Co-expression Network Analysis (MEGENA)^16^ and adapted the pipeline to produce novel analysis and plots. Application of permutation based false-discovery rate (FDR) estimates and building the network structures was extremely resource intensive, and for the present 5 networks, we implemented over forty trillion, three hundred sixteen billion calculations (**Supplementary Table 1**), with a cumulative 6-months+ of constant compute time. These five QNM represent valuable resources and are too extensive to detail in full here; instead, the utility of QNM to reveal novel human skeletal muscle biology is illustrated in the following sections by focusing on several examples. We will share a resource that presents all modules of each of the QNMs of human muscle in a browsable format (html link).

### Overview of the muscle quantitative network models

The bulk transcriptomic methods used assured representation from the non-coding genome on network architecture^15^ – and 37% of significant hub-genes common (n=104) to both young and old models were non protein coding genes (**Supplementary Table 2**). Two of the network models focused on age, built using 540 samples balanced for equal representation from men and women, with an average age above and below the mid-life ‘switch’^11^(**Table 1**). The models of young and old muscle were complemented by three models using paired samples and *change* in gene expression responding to atrophy or hypertrophy (**Supplementary Table 3**). Approximately 2% of genes acts as significant network hubs. Hub genes originated from across the genome, with the expected numbers from each chromosome, except for a modest over-representation from chromosome 9 (n=20) and the Y chromosome (n=4), given their relative size (**Supplementary Table 4**).

Eighty significant hub genes (27%) common to both young and old network models had established roles in muscle biology; however, of top 50 hub genes (45% protein coding), 80% had no studied role in muscle biology. For example, of the commonly *top* ranked protein coding genes e.g. CEP131, CROCC2, IFITM10 or MUC6, none have a studied role in human muscle, illustrating the power of the methodology to identify new biology. This included ARHGAP4, the single most connected gene in human muscle, present as a hub-gene all 5 QNMs (**Table 1**). Of the other network hub genes common to young and old muscle, 37 (13%) were regulated by *in vitro* low-dose Rapamycin treatment, largely in skeletal muscle rather than endothelial cells (**Supplementary Table 5**), despite the genes being expressed in both. The remainder of hub genes were short- and long-noncoding RNAs, mostly of unknown function. We cross-referenced each network with disease expression signatures^10–12^, as well as a senescence signature represented by 673 literature sourced genes^44–48^. These were projected across each network, in combination with a novel cell phenotyping model, enabling a detailed view of each network module within the human muscle (**Figure 2A-D**). This method contrasts with the original hierarchal network plot (**Figure S1B**) where phenotyped modules of interest cannot be traced across the network nor annotated, and we will provide both the network model data files and the new analysis code, which will enable the analysis of enrichment of gene-lists beyond those presented here.

Our novel cell type phenotyping method integrated tissue, bulk cultured single cell and scRNAseq profiles^31,49,50^ to enable us to characterise each MEGENA-based QNM in detail. For example, Dobner et al^51^ used scRNAseq to define a large variety of endothelial subtypes, however given the susceptibility to dropout (lack of genuine transcript detection) it is challenging to differentiate cell-specific marker genes from technical artefact. To address this problem, we used bulk methods to profile individual cell types and so limit false-positive scRNAseq cell-type specific genes. For example, when uncorrected, scRNAseq-driven models predicted that >10% of bulk muscle network modules represented smooth muscle cells, when in fact they represent <2% of the biopsy. Adjusting cell specificity scores with rank order bulk signal and the relative and absolute expression pattern bulk profiles of cultured primary cell types (e.g. myocytes, macrophages and T-Cells) helped limit rare cell-type specific classification rates. We find that the root-modules within the hierarchal network structure of human muscle are dominated as expected by signal from a mixture of cell types (e.g. **Figure 2A**), while the more distal branches are dominated by cell type specific modules indicating a degree of cell-type deconvolution. An example of a Type I fibre-type dominated hierarchal module structure is presented in **Figure S2**. He we find that a set of nested myocyte enriched modules, that reflect Type I slow oxidative muscle fibres. A key hub gene is SERCA2 (ATP2A2), the calcium pump protein isoform dominant in Type I fibres^52^ and these modules contains other genes that our spatial technologies and the literature confirm are enriched in human Type I fibres (See below).

While, as expected, myocyte-dominated modules were the most prevalent, we also identified fibroblast and endothelial dominated modules which – as discussed below – play a central role in the coordinate expression of senescence related genes. As we have previously noted^31^ we can identify immune cell co-expression modules, representing <1% of cells, in a muscle biopsy. The dynamic networks (during atrophy and hypertrophy) demonstrate a greater number of myocyte dominated modules (**Figure 2C, 2D** and **Figure S1C**) reflecting the load-detecting cells dominating the transcriptome. Nevertheless, as we show below, both positive and negative tissue remodelling intimately involves multiple cell types, especially including the vascular system. Each module will be explorable using our online resource (html link), where all the co-expression partners can be directly visualised, something not captured by snRNAseq or scRNAseq methods.

### Global DE analysis of the responses to muscle age and load status

Prior to introducing a machine-learning based explainable model of the transition from young to old skeletal muscle, we first introduce examples of in vivo and in vitro perturbation studies (i.e. altered muscle loading) and present key global characteristics. An overarching aim is to illustrate how network models compare with and provide novel insights beyond typical DE signatures when studying muscle age (**Figure 3A**). For the DE signatures we use >1,100 human samples, including comparisons of younger to older human muscle, frailty status, atrophy, hypertrophy, cardiorespiratory conditioning, sex and produce experimental signatures such as 100nM Rapamycin transcriptomes in muscle and endothelial cells, as well as an aggregated literature signatures for cellular senescence.

**Figure 3.**
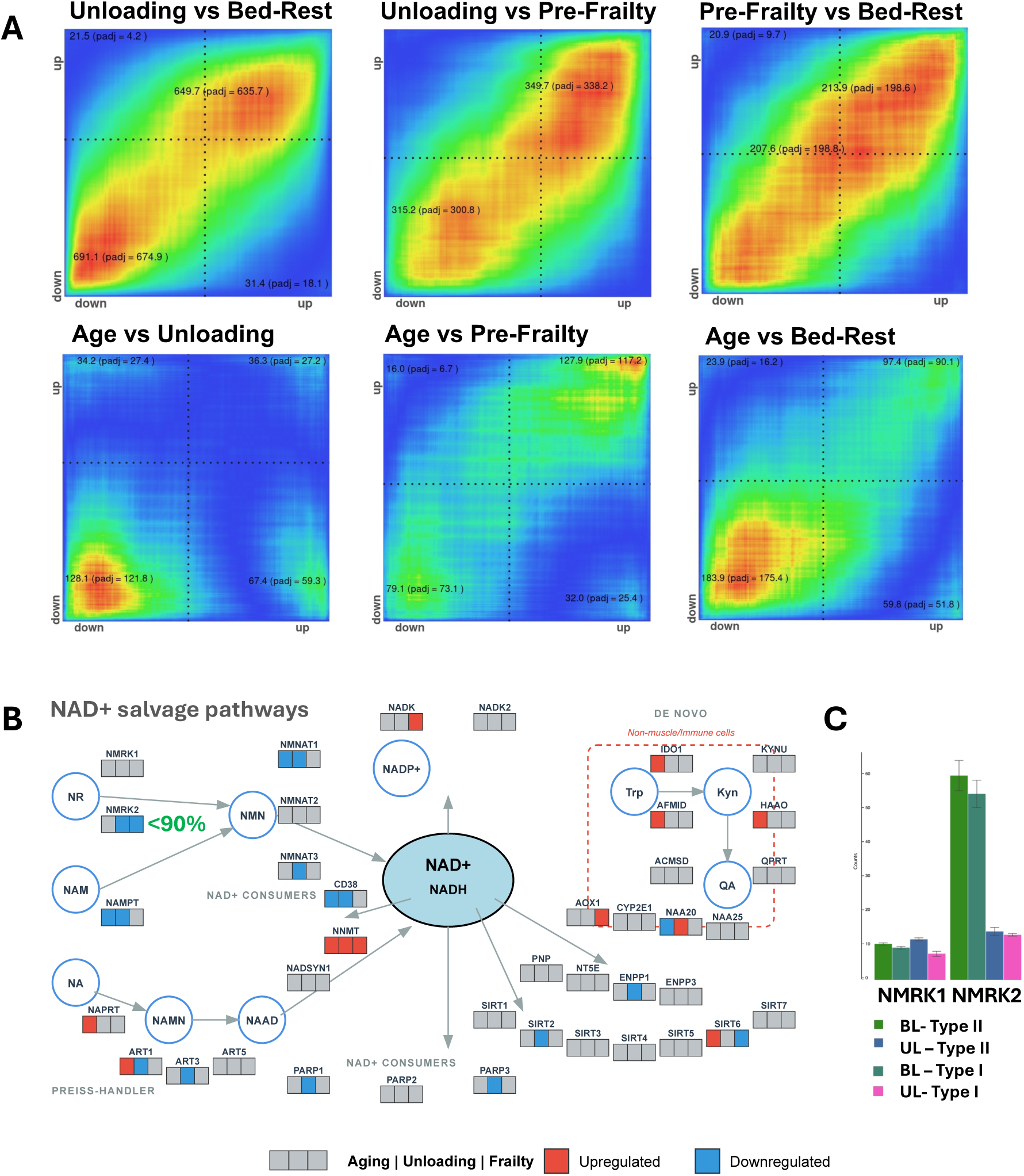
A) Hypergeometric rank analysis of fold-change values across 28,000 genes. Fold-change values are generated from young and old muscle (n=540) profiles balanced for sex. Bed rest profiles represent the re-worked analysis of the Meta-Predict FP7 funded profiling of X bed-rest samples (n=11) and the samples from van Loon et al reprofiled on the HTA platform (Age=x, n=76) plus muscle unloading in young and older individuals (n=47) B) NAD+ salvage pathway response by age, UL47 and frailty. C) Fibre specific expression of NMRK2 using the GeoMX spatial technology. NMRK1 is barely expressed while NMRK2 expression is lost in both fibres during unloading induced atrophy.

It is unclear if age-related frailty is a more rapid form of physiological aging or whether it is dominated by factors driven by reduced physical activity. Thousands of genes were DE across these comparisons, with 3,304 genes (1.25FC, <1%FDR) for young vs older muscle (**Supplementary Table 6**) and by using a rank-based comparison^53^ we can contrast conditions with distinct sample-sizes in a robust manner^53^. We report for the first time that both short-term (1-2wk) or longer-term bed-rest in younger healthy muscle share substantial commonality with a pre-frailty signature in muscle from ∼74y old individuals (**Figure 3A**). In contrast, muscle age DE (**Table 1**) has limited features in common with experimental disuse (e.g. loss of metabolic gene expression). This finding contradicts the claim made by Voisin et al^54^ where age and disuse were reported to strongly correlate (*Figure 3a* from Voisin et al^54^). It is noted that their association was generated from a linear correlation applied two distinct populations of values. Their muscle disuse DE signature was obtained from metamex^14^ and here muscle disuse studies are confounded by other interventions (e.g. GSE104999). We noticed that signal for some of the most robustly regulated ‘disuse’ genes e.g. RUNX1 or NMRK2, were not reliable represented in the Metamex^14^ data-base. In general, we find in our analysis that most of the global age-related DE gene changes presented were not mimicked by muscle disuse, and that pre-frailty is not simply a phenotype of accelerated age, findings that together cast serious doubt on the conclusions reached by Voisin et al^54^.

Aging is associated with reproducible defects in metabolism^55^, including multi-tissue coordinated loss of mitochondrial gene expression,^11^ which in turn is thought to lead to a greater risk of frailty^56^. A feature reported to be characteristic of aging is disruption of the nicotinamide adenine dinucleotide (NAD+) pool^57^. The nicotinamide riboside (NR) kinase (*NMRK1* and *NMRK2*) genes are the major salvage route for using exogenous NR to replenish the NAD+ pool^58^ and these metabolic processes are directly connected via MCART1 mediated transport^59^. In mice loss of NMRK2 and depletion of NAD+ are a feature of cachexia^60^. There seem to be some positive benefits of manipulating the NAD+ precursor pathways^61–63^ however under conditions of disease or muscle disuse it would be important that key salvage pathways are sustained, and we find that aging, frailty and unloading show overlapping systematic loss of key genes in this pathway (**Figure 3B**). In mice, NMRK2 is reported to be restricted to Type IIx muscle fibres^64^, however, we show in humans that NMRK2 equally expressed in Type I and II fibres and dramatically down-regulated during muscle disuse (**Figure 3C**). This discrepancy highlights a need for caution when attempting to identify the determinants of human metabolism, vis-à-vis using KO mice^65^, because of, among other things, compensatory mechanisms^66^.

Numerous other metabolic genes were DE with age and load, including more linked to NAD+ biochemistry. For example, Triosephosphate isomerase (TPI1, which we previously linked to muscle aging, works as a homodimer and catalyses the isomerization of glyceraldehyde 3-phosphate (G3P) and dihydroxyacetone phosphate (DHAP) in glycolysis and gluconeogenesis). Numerous lines of evidence indicate TPI1 protein has functions beyond glycolysis. This “moonlighting”^67^ by TPI1 illustrates the extreme challenges when trying to interpret DE, even for genes within well-established metabolic pathways. Network analysis reveals functional rewiring of OXPHOS processes (**Figure S3**). For example, SDHB represented a hub gene in young muscle, and becomes a member of a module with SUCLG1 (succinate-CoA ligase) as the hub in old muscle (**Figure S3B**). SUCLG1 promotes mitochondrial biogenesis via preventing the succinyl-CoA mediated succinylation mitochondrial polymerase POLRMT^68^ and it becomes far more connected in old muscle (Node.Strength=7.8, Node.Strength=80.6, and see below). It is plausible that the age-related loss of SDH capacity results in promotion of POLRMT succinylation (through succinate accumulation), leading to compensatory (but partly ineffective) promotion of SUCLG1 activity. Notably DE analysis does not show a measurable change in SUCLG1 gene expression (-1.07FC, n=540), highlighting the distinct view gained from QNM vs DE.

For our exercise training intervention studies, we noted the well-established non-responder phenomena^1,2,8,9,69^. In response to any type of exercise intervention, individual molecular responses diverge when contrasting responders (numerical majority) with non-responders (minority) – where responder status refers to a major physiological phenotype^7,9,70^. Until now, no reliable genome-wide analysis exists for the distinct DE profiles for non-responders for muscle hypertrophy or aerobic adaptation (**Figure S4**) due to a lack of sufficient non-responder profiles on a standard transcriptomics platform. As we demonstrate in **Figure S4A**, there are a subset of a few hundred consistently induced genes in hypertrophy responders and a small cluster of diametrically opposing responses. Likewise, for those that demonstrate an improvement in aerobic capacity (**Figure S4B**) there’s a common induction of extracellular matrix and vascular pathways irrespective of functional adaptation. Moreover, responders for hypertrophy and aerobic capacity show a DE pattern with more in common, particularly up-regulated genes (**Figure S4C**), than contrasting responders and non-responders for muscle growth. This helps explain why early attempts to carry out meta-analysis of human muscle transcriptomics data have limited validity^14^, containing false positive and negative data^24^ because no physiological context was considered when calculating mean changes in gene expression (the ‘gene’ signal also combined distinct genomic entities). The same defect applies to other online resources (e.g. https://www.extrameta.org/). In the present work, we use a consistent technical platform, identical genomic alignment and calculate mean-based values using people with comparable physiological responses. Further, it has been noted that these meta-analyses frequently exaggerate the sample sizes driving the key analyses. For example, Gene SMART^54^ is claimed to provide 66 individuals responding to exercise, when there are only 57 baseline samples and only 17 after 8wk training (severe unexplained drop-out; GSE171140). In general, published meta-analysis resources fail to highlight that key findings were often established in the original studies.

Melov and Tarnopolsky claimed that resistance exercise reversed muscle aging^71^ illustrated by highly selective presentation of DE gene responses. A global re-evaluation of that data finds no evidence for ‘age reversal’; quite the opposite, globally DE profiles between young and old muscle was similar when calculated using either pre or post exercise training samples (**Figure S4E**). We present a larger comparison of DE profiles versus a more robust (**Table 1, n=540**) and independent age DE profile (**Figure S5**). We find that resistance training of *old* (**Figure S5B**) but not young muscle (**Figure S5A**) induces a pattern consistent with the “reversal” narrative. Intriguingly both acute and chronic muscle unloading and pre-frailty appears to induce pathways that are potentially compensatory – attempts to protect muscle integrity – as small clusters of ‘atrophy’ related DE patterns are found in common with resistance or endurance training (**Figure S5D-K**). Further, long term unloading (∼ 3months bed-rest) induces clusters of gene DE which oppose that observed with endurance and resistance training (**Figure S5G-I**). Later we integrate these data sources with the QNM to produce a machine-learning based model of the transition from young to old muscle.

### Spatial, Single Cell and Bulk transcriptomic comparisons

We utilised the Merscope, Xenium and GeoMX platforms to further contextualize our findings^72–74^. The GeoMX methodology requires pre-staining of the individual cell types and the immunohistochemical (IHC) images are used to segment the muscle section *during* profiling where bar-coded oligos are collected from specific cell types, for off-machine sequencing (**Fig S6B**). The Merscope produces true single-cell high resolution images for multiplex subcellular transcript counting and assignment to a pixel level image of the muscle tissue^75,76^. Type I and II fibres are ‘stained’ using Fluorescent In Situ Hybridization (FISH) probes targeting high abundance specific genes (MYH7, MYH1 and ATP2A1) while other cell types are identified using merfish^77^ probes targeting known cell specific markers (See methods). This includes a recently identified novel nuclear expressed long non-coding RNA (MYREM, ENSG00000268518), that we identified marked mature human muscle nuclei^31^. The Xenium platform delivers somewhat similar data to Merscope, except fibre type marker RNAs are included within the main assay, as there is no auxiliary FISH channel for high abundance genes. Modelling of spatial transcriptomics data pose new^78^ challenges for data handling, quality control and application of deep learning approaches represent recent developments, and many studies have segmentation problems^79^ such that analysis of the Merscope and Xenium will require further development. Here we release the GeoMX data in full, and provide targeted analysis from Merscope and Xenium (while further data is generated, and segmentation models are built). The present release is the largest and most complete resource for studying the human skeletal muscle transcriptome, avoiding the cDNA library diversity limitations of Illumina short-read sequencing^6^.

We have demonstrated above that QNM can segregate out a robust Type I muscle fibre hierarchal cluster (**Figure S2**). We profiled 8 biopsy samples across 4 independent GeoMX slides, pre-staining the sections for nuclei (SYTO13), Type II fibres (MYH2) and endothelial cells (CD31). Individual fibres were selected by positive (Type II) or negative selection (Type I) and the oligos pooled for bar-code-based sequencing using the GeoMX work-flow^74^. The technology demonstrated no ability to yield gene expression from the pooled endothelial areas of interest (AOI) while the true single cell technologies can. In total 57 libraries (AOI’s) yielded gene expression, giving us technical replicates for normal loaded muscle samples^24^ (Samples - 1A, 4A, 8A, 9A, 11A, 12A, 13A and 14A) and a single profile of matching post-unloading samples (Samples 4C, 8C, 9C, 11C, 12C, 13C and 14C were intact), all of which have a matching HTA bulk profile.

As can be seen in **Figure 4A**, GeoMX based profiling produces fibre type specific gene expression and patterns consistent with independent dissected single-fibre proteomics^52^. In humans, the fraction of the tissue biopsy (the volume taken up by Type II fibres) reduces with age, as their cross-sectional area declines^4,80^. There is limited evidence that there is specific loss of Type II fibres^81^, nor is there clear evidence for a deficit in satellite cell number in aging human muscle, at least in the context of being able to respond to changes in load^82–84^. However, anatomical factors such as proximity to vascular cells and other factors regulating the satellite cell niche are potentially important^82–84^ and these relationships can be modelled using QNM approach models or directly visualised using spatial transcriptomics. Because the GeoMX platform was unable to produce endothelial profiles, we focus on the analysis of human muscle fibre type specific responses before and after atrophy induced unloading (**Figure 4B**), noting genes (red-arrows) that feature in the QNM analyses.

**Figure 4.**
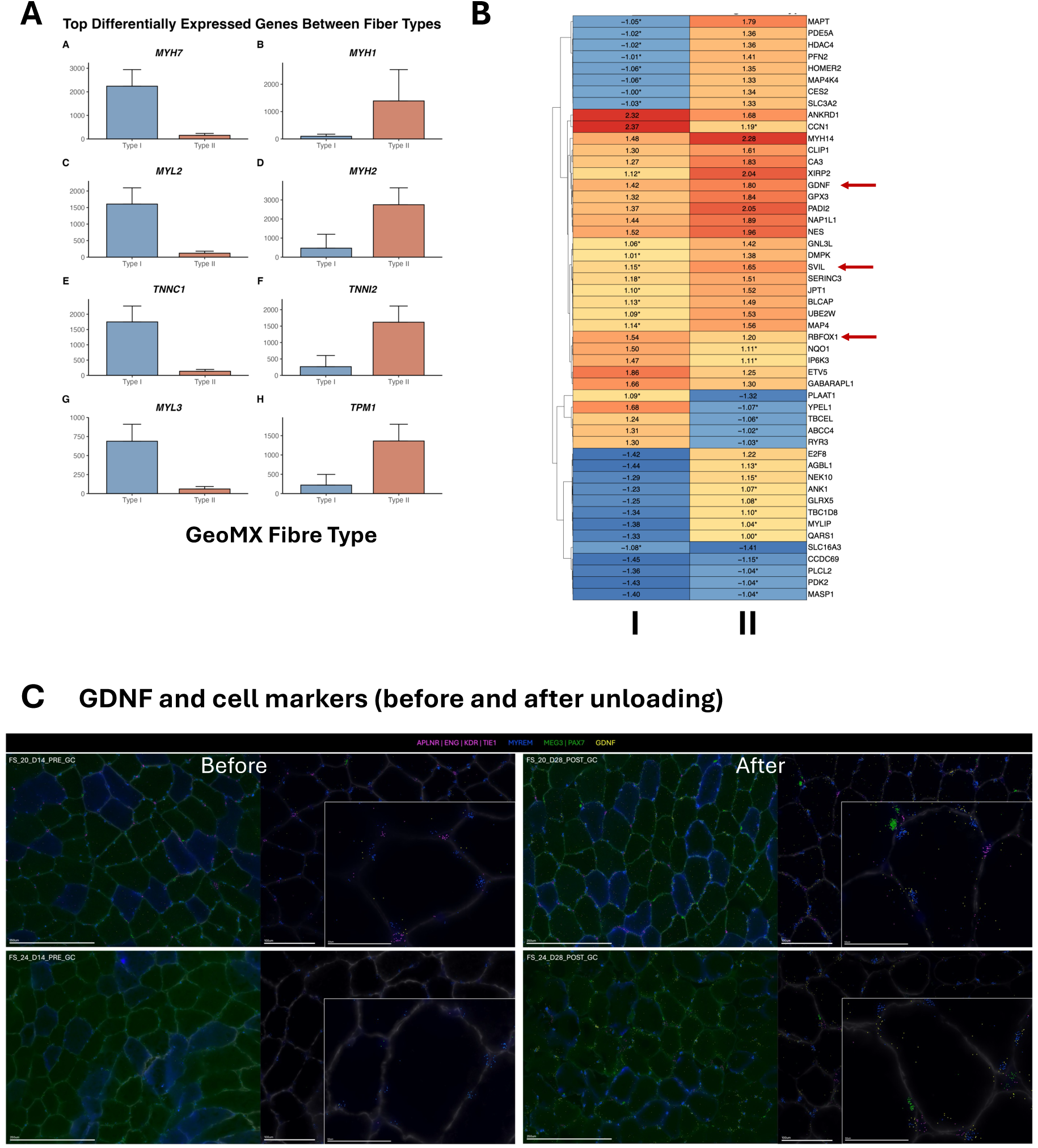
A) Fibre type specific gene expression in agreement with Murgia et al 2021. B) Using GeoMX we find a number of genes are differentially regulated in Type I vs Type II fibres, with atrophy – this includes three network module genes, GDNF-SVIL-RBFOX1 C) Merscope profiles before and after human muscle unloading. GDNF (yellow) expression in four ROIs (before and after 2 weeks of disuse). Mature muscle nuclei (MYREM+; blue) muscle satellite cells (MEG3/PAX7+;green) and endothelial cells (APLNR+/ENG+/KDR+/TIE1+; pink). At low magnification fast (ATP2A1, green) and slow muscle (MYH7, blue) fibres are stained. At higher magnification, fast are unstained (ATP2A1 fish channel switched off) and slow fibres are blue (MYH7). Clear examples of GDNF expression in or around mature skeletal muscle nuclei (MYREM+; blue) are show with 4-10 times more GDNF after unloading and consistent with the bulk analyses (Supplemental Table S7)

Over the past two decades, numerous reviews have labelled cytokines as muscle secreted factors. Mostly, there is no direct evidence for *in vivo* muscle cell expression of the genes in question, while arterio-venous differences in blood concentration (which is then multiplied by estimated blood flow to report net secretion) can reflect a multitude of factors, including circulating cell activation, ‘wash-out’ of post venule immune cells, and water movement across compartments. We characterised some popular muscle associated cytokines using GeoMX, Xenium and Merscope technologies. Unlike prior studies, which relied on aggregated signal from cells in a muscle biopsy and qPCR^85^ (which is usually house-keeper scaled and so ∼ abundance is obscured), spatial technologies measure expression directly *in situ* with precise single molecule resolution.

We profiled 60 biopsy samples (‘Regions of Interest’, ROI) on the Merscope and 54 ROIs were physically intact after visual QC (**Figure S7A**). Serial sections, on distinct slides (∼technical replicates), yield correlations for merFISH detected gene expression of ∼ R=0.9 (**Figure S8B**) demonstrating good reproducibility. Merscope auxiliary FISH based marker gene analysis of Type I, II and IIx fibres confirmed two things – human muscle robustly expresses MYH1 (IIx), mostly co-expressed with ATP2A1, and thus represent hybrid Type II and IIx fibres as described by Schiaffino’s lab^52^. This means that our GeoMX type II fibre data is almost certainly hybrid signal from more than one type of “Type II” muscle cell type (**Figure 4A** clearly shows both MYH2 and MYH1 enrichment). One advantage of the Merscope is that we can examine individual fibre gene expression to confirm that absence of e.g. MYH1 signal or apply modelling agnostic to any classic muscle fibre type designation. Using both the GeoMX and Merscope, we show that Glial cell line– derived neurotrophic factor (GDNF) gene expression is induced with unloading (**Figure 4B and Figure 4C** – zoom-in to view individual GDNF RNA molecules in high-res image). Both bulk (2,546 genes DE, 1.25FC, <1FDR, **Supplementary Table 7**) and GeoMX identified substantial DE after 1-2wk muscle unloading (**Figure S6C**).

GDNF is one the most up-regulated genes in the bulk analysis with unloading (**Supplementary Table 7**). GDNF represents a genuine muscle secreted protein^86^ with a local paracrine role, and somewhat ironically it is induced by muscle inactivity, not exercise (**Figure 4C** – Panel inserts Zoom in to view these high-resolution images). GDNF plays a crucial role in early motor neuron development, as well as neuromuscular junction maintenance and muscle innervation in adulthood, acting through RET/GFRα1 activation^87^. GDNF appears to promote axonal outgrowth, and enables muscle reinnervation after injury, ultimately restoring neuromuscular function^88^. Unlike GDF15, which binds to a GDNF-related receptor^89^, a broader physiological function for GDNF is unclear, and here we produce evidence that in the first weeks of muscle unloading it is robustly expressed in all muscle fibres, not just proximal to neuromuscular junctions (GDNF is in a myocte dominated module during atrophy (UL47_ c1_937) and also co-expressed with the aging, IR^11,12^ and splicing gene RBFOX1, and the myopathy linked gene SVIL (html link). We find these three co-expressed genes are modulated somewhat distinctly in Type II muscle fibres (**Figure 4B**-red-arrows) using the GeoMX spatial technology, with GDNF and SVIL increased more and RBFOX1 less so (the potentially limited dynamic range of the GeoMX becomes apparent, as these genes are 4-,1.6- and 1.7 fold up-regulated in the bulk data, **Supplemental Table 7**).

Our bulk data analysis indicates that after a couple of months of bed-rest the muscle no-longer mounts an increase in GDNF expression - whether this reflects a slowing down in the rate of muscle atrophy, or some feedback that enhanced GDNF signalling to compensate for the lack of muscle use, is no longer working, is unknown. Lai et al^21^ reported a linkage disequilibrium analysis integrating snATACseq profiles with a previous GWAS study on muscle weakness (that reported 15 genome wide associations) and other anatomical phenotypes. Lai et al^21^ reported that lean body mass variants were enriched in Type II nuclei expressed genes, while muscle-strength genetic traits were linked to fibroblast-like and fibro-adipogenic progenitors (FAPs) gene expression, but not in myofibres (yet we know the transcriptomic resource will suffer from GE drop-out). Variants associated with lean mass and grip-strength identifying 3,158 candidate variants, including a GDNF locus in myofibres (rs1862574), adding support to our spatial analysis of GDNF response to unloading induced atrophy, and the decline in GDNF expression with age.

There are numerous reviews^90^ claiming that human muscle is a major secretory organ especially during exercise, where secretions are claimed to contribute to the systemic effects of routine physical activity. Many cases include low absolute levels of cytokines (e.g. IL6), possibly reflecting release from tissue resident or blood borne immune cells. Alternatively, local production of these factors in human muscle may reflect a role in a local autocrine or paracrine action within skeletal muscle biology, with blood spill over being coincidental. It is challenging to assign a physiological role to these cytokines in vivo in humans. For example, recently reported marginal effects of IL6 on fatty acid metabolism^91^ used tocilizumab, an antibody that targets the IL6 receptor, not IL6 protein. The receptor, IL6R, partners with gp130 (a more general receptor partner), and tocilizumab induces IL6 receptor expression itself, altering gp130 partnering stoichiometry with unpredictable impact on other gp130 partners^92^. In this study the metabolic differences assigned to IL6 likely reflect the fact that the tocilizumab group did 12% more work (due to using fraction of VO2max and not work).

Do these ‘muscle tissue’ secreted cytokines originate from muscle cells and hence merit a ‘myokine’ label? IL6, the most cited exercise ‘myokine’, has only one study that made a direct determination of its mRNA and protein inside human muscle fibres^93^. The Febbraio lab used the now discontinued rabbit polyclonal IL-6 antibody at high titre (1:20, H-183, Santa Cruz Biotechnology; replaced with more specific monoclonal reagents since). It is also unclear if the image acquisition was appropriately gated as no secondary-only control sections were presented. The single post-exercise example had a signal dominated by the vascular compartment (which could be blood sourced or reflect endothelial cell IL6 production). They report that Texas red and biotinylated streptavidin-peroxidase gave non-specific binding, while fluorescein did not, however given 63 sections were processed it is unfortunate that the article provides only one example of exercise induced fluorescein-based protein staining.

The same study reported an 18-fold increase in IL6 mRNA using a 18s qPCR house-keeper approach and a radio-labelled nucleic acid probe targeting a 1.23 kb human sequence from GRCh37 (hg19). Substantial pre-exercise IL6 mRNA expression is visually presented in the one ISH example given. The pattern of IL6 RNA detection was peripheral, just as we see for hundreds of genes in muscle fibres using the Merscope. Post exercise the Febbraio lab^93^ report IL6 mRNA signal was now predominantly in fibres rich in glycogen – a molecule known to trap RNA^94^. Biophysical interaction with glycogen may explain why the RNA signal came largely from the remaining glycogen-rich fibres, as the other fibres were now depleted of glycogen through exercise. Access to the remaining ∼59 sections would clarify how representative these data are. Notably, since the Febbraio lab^93^ study, the IL6 loci has been revised in detail and location (from Chr7: 22,765,503–22,771,621 to Chr7: 22,725,889–22,732,002) so it is unclear if the (now replaced) polyclonal antibody, or the radio-labelled nucleic acid probe are specific for IL6. Thus, to date there remains to date no reliable evidence for IL6 RNA or protein expression inside human muscle cells, in vivo.

Using the Merscope we directly imaged individual IL6 mRNA molecules (relying on >50 specific ∼27-mer probes), along with images of the IL6 receptors, in loaded and unloaded human skeletal muscle. The number of IL6 mRNA molecules detected across a muscle biopsy was no different from the level of signal from the background non-specific control probes (∼500-800 per ROI). However, the single-cell pattern of gene expression also needs to be considered. Interestingly, we found very rare examples (**Figure S8**) of robust endothelial specific intracellular IL6 mRNA expression (unrelated to load status), with more IL6R mRNA molecules in unloaded muscle - in this single example. Interestingly, unloading healthy muscle is known to induce insulin resistance^95^. These endothelial images demonstrate the Merscope IL6 assay is working and across the rest of each muscle section (ROI), we observe single IL6 mRNA molecules in every 10 or 20 fibres (indistinguishable from the negative control probes). We present images from two of the examples with the most IL6 counts we could find, one with and the other without loading (**Figure S8**). Note the pattern of IL6 signal inside muscle cells was no different from areas of slide with no tissue (**Figure S9**). We then used an entirely different technology (Xenium technology) and again find very rarer examples of a single IL6 mRNA molecule inside a muscle cell (**Figure S10**).

Thus, other than in rare ‘inflamed’ endothelial cells, human muscle does not reliably express IL6 under chronic load or unloading. This probably helps explain some reported extreme fold-change values using qPCR following acute exercise (an extreme delta-CT value being driven by dividing by a near zero value). Applying the Febbraio labs^93^ reported 18-fold relative increase in IL6 would represent a single molecule in each muscle fibre (assuming no contributions from non-muscle cells e.g. activated immune cells and endothelial cells, which seems extremely unlikely). If we utilise a meta-analysis of acute human exercise muscle transcriptomic profiles^14^ and analyse cell-type matches^96^ with genes co-expressed (R=0.86-0.67) with IL6 (which is ∼5 fold DE, n=295) and then analyse which cell types this signature represents, we find that the cell types represented by the top 100 covarying IL6 genes in muscle tissue are monocytes, macrophages and endothelial cells (<1×10^-20^, **Supplemental Table 8**) further supporting the idea that even when IL6 is induced by exercise (from zero to minor levels) it is not in muscle cells.

Spatial transcriptomics is a technology that is potentially better suited for studying multi-nucleated fibrous skeletal muscle than scRNAseq and snRNAseq. Recently there have been two human muscle atlas projects using scRNAseq and snRNAseq to study age. In the first study, Kedlian et al^18^ produced scRNAseq and snRNAseq analysis of human intercostal muscle age - relying on 17 donors of distinct ages reporting observations on senescence, denervation, inflammation and metabolic dysfunction (all factors influenced by muscle load status^3–5^). Kedlian et al^18^ profiled ∼90K cells with each method while the anatomic interpretation of the nuclei used *MYH1, MYH2* and *MYH7* marker genes. Kedlian et al^18^ reported that only 4.7% of human muscle “age genes” were DE in mouse skeletal muscle. They found that most cell types display a pre-ponderance of down-regulated genes with age, with only inflammation related processes showing up-regulation. However, this was based on GTEx data, where both disease and use of RNAseq^6^ limits its utility for studying muscle or aging. We report, using a data set >30 times larger and far less biased transcriptomic profiling^6^, that >3,300 genes are regulated, up and down in equal amounts, representing a broad biological narrative.

In a second muscle study, Lai *et al* presented a cell atlas of human muscle, with particular emphasis on age related changes, using tissue from 31 people^21^. They reported using human ‘hindlimb’ muscle (17 men and 14 women), covering two discrete age-ranges (15-46y, n=12 and 74-99y, n=19). Thus, any age-related differences reported may reflect muscle regional anatomy (and sex). The histological profiles, snRNA-seq, snTAC-seq and scRNA-seq data came from a subset of the 31 people (∼7 for young, and up to n=15 for old). While n=3 and n=4 was used to estimate overall cell composition per biopsy. The analysis reflects scRNAseq profiles from ∼80K cells (∼2K cells per biopsy) and ∼220K nuclei and they found substantial variation in cell type by sex. The muscle studied was not just ‘hindlimb’ but *pectoralis major*, *gracilis*, *biceps brachii*, *vastus lateralis* and *gluteus medius* and each region was distinctly represented in each age group.

As snRNAseq and scRNAseq does not provide direct anatomical context, each nuclei is computationally assigned to a particular muscle fibre type. Lai et al^21^ did this based on the proteomics profile described by Murgia et al ^52^ such that nuclei from three main fibres types were reconstructed: Type I, Type II and Type IIx. The typical Type I marker gene is MYH7, ATP2A1 or MYH2 for Type II, and MYH1 for Type IIx. Type II fibres are often hybrid expressing both ATP2A1 (or MYH2) and MYH1. We also find individuals where MYH1 appears dominant in most Type II fibres with little evidence that ATP2A1 was co-expressed (**Figure S11A-11I**). The belief that muscle nuclei only express a single fibre type myosin heavy chain isoform related to its host muscle fibre^97^ is inconsistent with in-depth analyses using lineage tracing ^20^.

We show here, using the Xenium spatial transcriptomics platform, that muscle nuclei located in fast muscle fibres express both slow (MYH7) and fast gene (ATP2A1) markers and *vice versa* in slow Type I muscle fibres (**Figure S12**). This confirms the lineage tracing studies^20^ and indicates these genes cannot be used to pool snRNA-seq data from skeletal muscle in a fibre specific manner. It is unclear which other genes, selected from studies on muscle fibres, remain distinct in the nuclei meaning there is currently no ground truth for linking snRNAseq profiles back to the anatomy of muscle fibres. Variation in marker gene expression in snRNA-seq data reflects the high susceptibility for drop-out, such that subsequent differences that emerge, will be in part random. Notably Lai et al^21^ report a decrease in myonuclei with age, especially in Type II fibres – but here we have argued it is not possible to scale their data back to the level of the myofibril, as they have done, to make a valid calculation. In fact, it is known that as a fraction of the tissue biopsy, the volume taken up by Type II fibres reduces with age, as the cross-sectional area declines^4,80^.

Independent of anatomical fibre type discussions, Lai et al^21^ reported several age correlated genes. They noted that PAX7 (negative), PDGRFa (positive) and a general decline in clock genes PER1, PER2 and RORA in the nuclei. Relying on a data set 100 times larger, balanced for sex and using a consistent muscle type, we found the clock genes were up-regulated with age (PER1 (1.48FC, FDR<1%), PER2 (1.3FC, FDR<1%), RORA (1.17, FDR<1%) while BMAL1 was down-regulated (-1.19FC, FDR<1%) indicating that differences they ascribe to age may reflect other variables in their study. Notably their snATAC-seq data did not support their CLOCK gene observations. Lai et al^21^ stated that PCDHGA1^21^ was positively correlated with age “like our earlier work on frailty” (this is not correct as we reported on PCDHGA10 and PCDHGA11^98^, and both had a negative coefficient). In the present analysis PCDHGA1 was upregulated by age in our analysis, but it was not changed by unloading or frailty status, dissociating it from muscle functional status.

Lai et al^21^ reported AMPD3 was positively correlated with age, and in our analysis AMPD3 was dramatically up-regulated by various types of unloading but unrelated to either age or frailty. They found that the pro-atrophy activin pathway genes (ACVR1, ACVR1B, ACVR2A, ACVR2B) to be activated in aged myonuclei, with the ligand INHBB upregulated in non-muscle nuclei. We find that ACVR1 is modestly down-regulated (-1.3FC, 1%FDR, ACVR1B (ns), ACVR2A (ns)) with only ACVR2B being up-regulated with age (1.2FC, 1%FDR) albeit in bulk muscle tissue. None were altered by muscle unloading or frailty status. In general, most of the examples provided by Lai et al can be challenged and this undoubtably reflects the use of a small sample size, the ad hoc mix of muscle types and the distribution of sex in their small cohort^21^ (as well as limitations of the scRNA-seq and snRNA-seq methodologies applied to multi-nucleated skeletal muscle). Notably, the Pearson correlations reported by Lai et al^21^ were driven by two entirely discrete data clusters (due to the discrete age range of the samples used) while regardless, small samples typically render correlations unstable^41^.

In short, our data challenge most of the detailed ‘ageing’ examples provided by Lai et al^21^. Instead, we present a network driven evaluation of aging and senescence genes in ∼28,000 genes measured in whole muscle below, isolating cell type contributions using a novel enrichment model (that combines scRNAseq and cell culture data). We then go on to present a machine-learning model of the network topological changes found between 270 young and 270 older people, one that highlights that muscle ageing is strongly connected to network features of long-term atrophy, rapamycin signalling and insulin resistance.

### Senescence-related myocyte and satellite cell modules in older human muscle

It is believed that muscle atrophy is partly mediated by rarer cell types in skeletal muscle, including endothelial cells and fibroblasts. However, the underlying gene expression programs regulated by these cell types may be obscured/minimized in DE analysis; therefore, we reasoned that QNM would provide greater insight into these relationships and here we present several modules containing genes predominantly enriched in these rare cell types. We carried out an in-depth human-driven analysis of several QNM modules, in a manner blinded from the results of the machine learning modelling. While traditionally associated with mitotically-active cells, senescence pathways are increasingly recognized for their roles in growth regulation and homeostasis in terminally-differentiated tissues,^99^ which may therefore include myofibers.^100^ Alternatively, resident mononuclear cells (e.g., fibro-adipogenic progenitors (FAPs)) have emerged as key drivers of senescence in skeletal muscle in response to injury and aging,^100,101^ yet whether these processes appear distinguishable in unloading-induced atrophy in healthy skeletal muscle is unclear. Moiseeva et al.^101^ employed an innovative fluorescence-activated cell sorting approach in which SA-β-gal positive cells were selectively labelled to isolate populations enriched for senescence markers. Using this approach, they found that monocytes, macrophages, FAPs, and satellite cells (SCs) constitute the primary cell types in skeletal muscle prone to senescence^101^. A longstanding question in human aging is whether altered satellite cells play a role in muscle aging, in a manner like mice^102^. Kedlian report that satellite cells show an increase in CCL2 with age, but this was not evident in bulk vastus lateralis. Instead, we find that short term muscle unloading, increased CCL2 (1.7FC, FDR<1%), as part of a CD63 (Grey) tetraspanin coordinated module enriched in senescence and insulin resistance genes (**Figure S13**). During atrophy, CCL2 was directly connected to KPNA2, which is negatively associated with insulin sensitivity^12^ and ENO1, a senescence and neuromuscular age correlated gene^11^.

Our QNM analysis identified several senescence-enriched modules, which had cell type profiles including macrophages, fibroblasts/FAPs, satellite cells, and/or endothelial cells. This included several Senescence Associated Secretory Phenotype (SASP) factors identified by Moiseeva et. al^101^ – such as *TIMP2, IGFBP4, CCL2*, and *CCL7* (html resource). The classic senescence genes p21/*CDKN1A* and p16/*CDKN2A* have low expression levels in human skeletal muscle being “nearly undetectable” by qPCR^103^. Attempts to overcome the rarity of expression has relied on complex sorting and enrichment procedures, as well as orthogonal experimental methodologies to identify senescence cells or expression signatures. For example, when Dobner et al^51^ adjusted for “tissue-specific transcriptional variation”, the anatomical (vascular tree hierarchy) pattern of endothelial cell specialisation became more apparent, supporting the idea that scRNAseq can be strongly influenced by non-biological variation. Contrasting an older group (63.8y mean age), with a younger group (35y mean age) Dobner et al^51^ found that there was reduced angiogenic capillary endothelial cells and that endothelial cells obtained from older muscle were enriched for senescence gene sets; although they still found p16/CDKN2A undetectable, while p21 (CDKN1A) was detectable and endothelial cells did not show enrichment for these genes. In vitro induced senescence in endothelial HUVEC cells yielded a candidate list of 331 genes, of which 153 were associated (enriched) in p21+ endothelial cells ex vivo. Using spatial methodologies, we show evidence that key senescence genes were expressed in skeletal myocyte dominated modules (**Figure 5**). P21/CDKN1A was four-fold induced during muscle unloading (**Figure 5B**) but not apparently DE regulated by age (**Figure 5C**). We also found that p16/CDKN2A was DE (up-regulated) specifically in Type I muscle fibres during atrophy using the GeoMX spatial platform, illustrating that spatial transcriptomics is likely to add further resolution to the study of senescence in human skeletal muscle.

**Figure 5.**
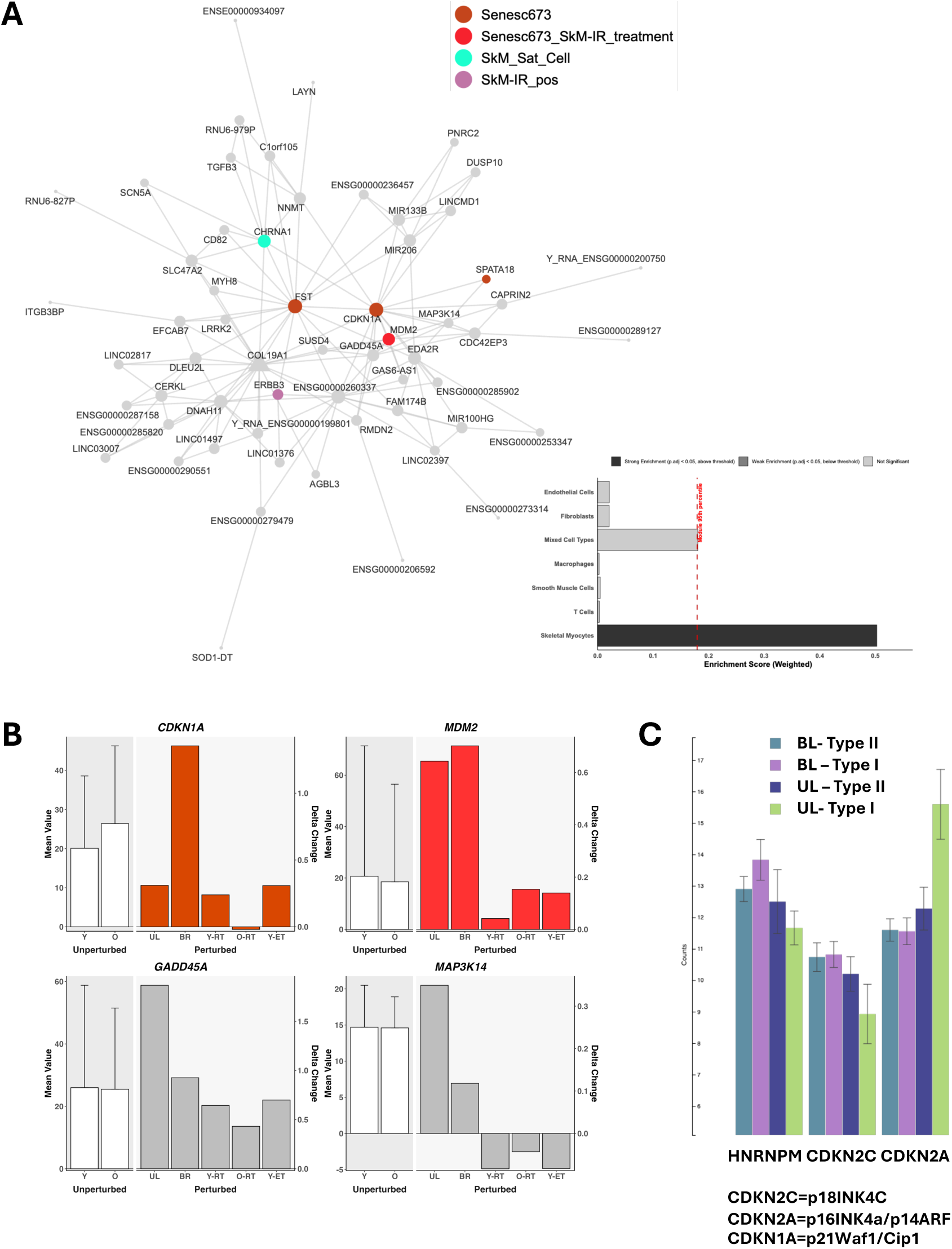
A QNM module identified from 28K genes identified a **c**lassic senescence marker gene enriched module within skeletal myocytes (Module c1_40_O270)

In addition to the potential influence of senescence the module in **Figure 5** also contains several genes with established roles in skeletal muscle atrophy. For example, *GADD45A* encodes a stress inducible protein induces skeletal muscle atrophy in a myriad of catabolic contexts^104^. Expression of *GADD45A* is low and in mice appears enriched in muscle satellite cells (https://tabula-muris.sf.czbiohub.org/visualizations), consistent with the idea that this module represents rare senescent cell populations. GADD45A is a myonuclear protein that influences myonuclear remodeling,^104^ particularly in type II muscle fibres, and is associated with muscle weakness in older adults^105^ and it may also serve a protective role in the setting of denervation-induced skeletal muscle atrophy. In addition to *GADD45A, CDKN1A* was directly linked to *MAP3K14* and *EDA2R* and *EDA2R* was recently shown to be in intimately linked with cancer-induced cachexia and to directly promote skeletal muscle atrophy via *MAPK314/NIK* signaling^106^

In older muscle, we identified a skeletal myocyte module with well-established senescence drivers, including *CDKN1A* (p21), *GADD45A*, and *MDM2*. While our cell-type deconvolution approach broadly pointed to skeletal myocytes as the enriched cell type, the gene composition of this network suggested that the signal was potentially from satellite cells (which is not a distinct category in our model). This is based on several observations. We note low expression levels of *CDKN1A, GADD45A,* and *MAP3K14* in bulk skeletal muscle and that can indicate gene expression from rarer cell types^31^. The module contains several genes enriched in myogenic stem cells/satellite cells, including *ERBB3, CD82* ^107,108^. Furthermore, a recent snRNAseq study identified a rare senescent satellite cell population, co-expressing *CDKN1A/MYH8/LRRK2/COL19A1/EDA2R*^109^, consistent with a similar senescent cell population in old myofibers^100^. Each of these genes was identified as an interconnected node in the old-muscle QNM (**Figure 5**). These findings underscore the utility of data-driven network analysis to uncover gene co-expression patterns arising from rare cell populations.

Having identified examples of rare senescent satellite cell population modules, we were interested in understanding its relation to age-related skeletal muscle dysfunction. While several gene members are known regulators of p53 signalling, other important connections emerged. For instance, key senescence genes (e.g., *CDKN1A, FST, and MDM2*) were co-regulated with canonical muscle-specific microRNA genes, *MIR206* and *MIR133B*^70,110^. These microRNAs regulate satellite cell differentiation and fusion^111^, and are correlated with sarcopenia^112^. Interestingly, miR-206 also plays an integral role in the reinnervation of skeletal muscle through regulation of bidirectional signalling between muscle fibre and motor neurons, a function central to Amyotrophic lateral sclerosis (ALS) progression^113^. Consistent with this role, we show that *MIR206* is a close network neighbours of *CHRNA1* and *FST* – both key neuromuscular junction genes^100^. Regulation of denervation-reinnervation cycles, observed in aged skeletal muscle, may lead to the accumulation of denervated and dysfunctional muscle fibres in older adult muscle^114,115^.

### Senescence-related endothelial modules in older and atrophying human muscle

In addition to myofibre-specific protein loss, skeletal muscle atrophy involves capillary rarefaction and remodelling of the extracellular matrix. Sensitive bulk transcriptomics coupled with QNM can reveal rare cell type processes^31^ with greater resolution due to the reliance of a more complete capture of the interacting genes. To identify modules from the QNMs enriched in senescence genes, we consolidated literature-derived senescence gene signatures into a single list, which was subsequently filtered to include only genes present in at least 2 of the 5 sources (resulting in n=627 senescence-associated gene, of which ∼480 were found in muscle, **Supplementary Table 9**). Next, each module in the QNMs was tested for over-representation to identify modules enriched in senescence genes. We found several senescence-enriched gene networks in aged skeletal muscle (e.g. **Figure 6A**) including endothelial cell dominated modules (**Figure 6B**). This module from the old QNM (n=270) included numerous vascular integrity and remodelling genes, a process disrupted upstream of anabolic signalling in older skeletal muscle^116^ and implicated in cellular senescence with advancing age^117^.

**Figure 6.**
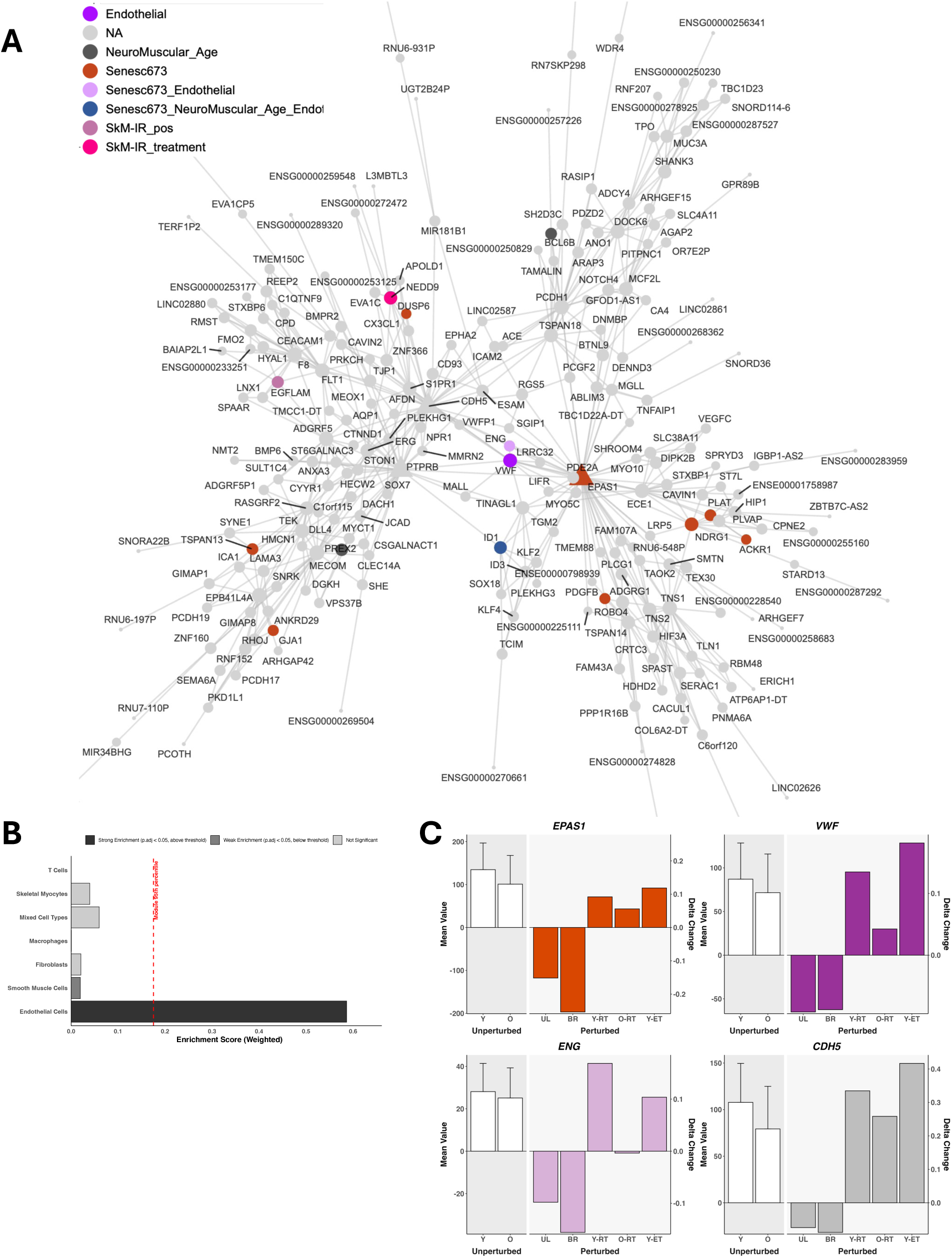
A QNM module identified from 28K genes identified an in vivo endothelial age-related senescence network module in older skeletal muscle (c1_153_ O270).

One of the hub genes in this module is the well-known oxygen-responsive transcription factor, *EPAS1*, which encodes Endothelial PAS domain protein 1 and is co-regulated with several genes enriched in endothelial cells (e.g., *LRRC32, AQP1, MMRN2, CLEC14A,* and *CYYR1*). Accompanying bar plots for selected genes (*EPAS1, VWF, ENG, CDH5*) illustrate their expression patterns with age, and relative changes across different load states **Figure 6C**. Another hub gene, *PTPRB,* encodes a tyrosine phosphatase that dephosphorylates two of its nearest network neighbours – *TEK/TIE2* and *CDH5* – in turn regulating endothelial cell proliferation, cell-cell contact integrity, and vascular remodeling^118–120^. *MMRN2* is directly connected to *PTPRB* and it is a known endothelial-cell specific extracellular matrix protein intimately linked with angiogenic signaling^121^. Further, most of the immediate neighbours of *MMRN2*, have been identified in endothelial single-cell sequencing datasets e.g., *ARHGEF15, ADGRL4, VWF, CLEC14A, CYYR1, NOTCH4, SPAAR, BTNL9, SOX18, CDH5, MEOX1, and PTPRB*) and we show they are directly connected to *MMRN2* or linked by a single connecting gene. MMRN2 directly interacts with CLEC14A and CD93 to regulate angiogenesis,^122^ reinforcing that transcriptomic co-expression relationships are consistent with protein level function. Key genes in this network were found to be downregulated in skeletal muscle of older adults following muscle atrophy (**Figure 6C**) indicating that unloading impairs vascular integrity, which has the potential to exacerbate pre-existing endothelial dysfunction in aged muscle in a load dependent manner^123,124^. Taken together, the network modelling reveals senescence-associated gene networks in vivo. The intra module-level interactions can be browsed online (html resource) provide unrivalled detail that can be used to design mechanistic studies.

We also produced QNM using *differential* expression data (**Table 1**), which illustrate modules of *coordinated* regulation during muscle atrophy. These analyses also revealed senescence-linked modules and again QNM revealed patterns of non-myogenic cell signatures, obscured in standard bulk DE analyses. For example, the SHANK3 and ERG coordinated module shown in **Figure 7** was enriched in genes predominantly expressed in endothelial cells. *ERG* encodes a transcription factor essential for endothelial cell homeostasis^125^ acting via other well-known regulators of vascular remodelling and homeostasis, including *VWF, DLL4,* and NOTCH^126,127^, signalling – all of which are tightly differential expressed during atrophy. In turn, they are connected to *EPAS1/*HIF2A which regulates cellular senescence^128^. Given the centrality of this gene network in endothelial cell homeostasis several hypotheses can be generated as to their specific roles during skeletal muscle atrophy. A recent pre-clinical study found that atrophy stimulated the production of the Notch ligand DLL4, which can trigger atrophy of skeletal muscle fibres^129^. We show how DLL4 is part of a module that in vivo responds to age and load, and more generally it is now appreciated that oxygen sensing factors, like *EPAS1* play a role in determining muscle growth and atrophy through coordinated vascular remodelling^31^.

**Figure 7.**
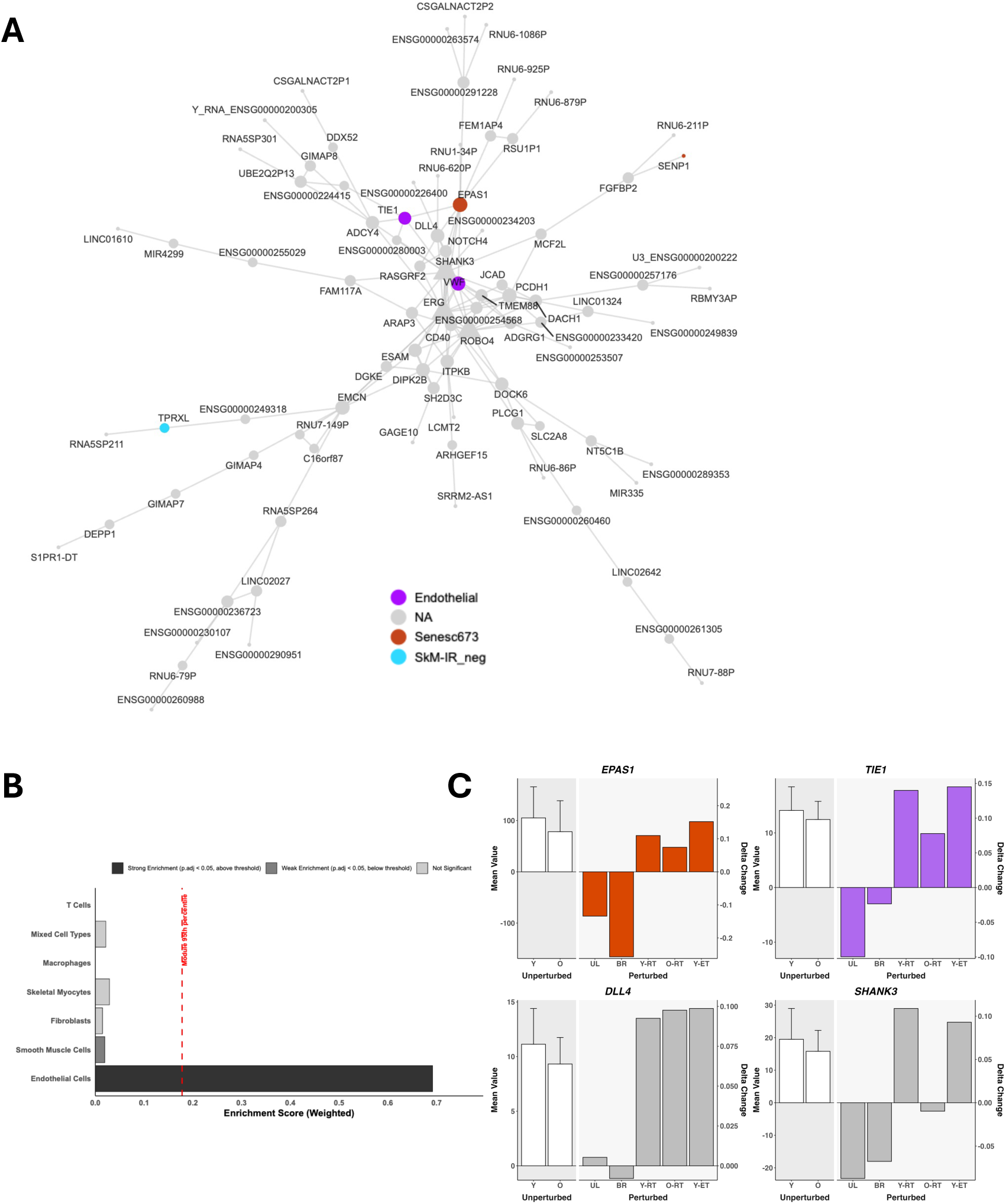
A QNM module identified from 28K genes identified a dynamic in vivo endothelial enriched network module from unloaded skeletal muscle (c1_140_ UL47)

Taken together, our findings extend or challenge recent single-cell studies of aging muscle. Moiseeva *et al*.^101^ demonstrated that senescent cells establish an aged, inflammatory niche that impairs regeneration in mice, while Zhang *et al.*^100^ identified distinct senescent cell populations within FAP progenitors and myofiber compartments. Our QNM based analysis of bulk human transcriptome points to senescence-associated modules within both the endothelial and satellite cell niches, as well as modulation of senescence genes in single muscle fibres, supporting the concept of multiple senescent compartments contributing to impaired tissue regenerative capacity or aging. Importantly, whereas Moiseeva and Zhang relied on single-cell isolation approaches to resolve these rare populations, our QNM approach extracts comparable insights directly from bulk co-expression network analysis where we have a more complete view of the transcriptome and greater representation from diverse human samples. Not only does this highlight the complementary nature of our bulk-transcriptomic QNM resources for studying skeletal muscle aging, it also illustrates the advantages of robust QNM methods over computationally light-weight correlative methods^130^ or classic DE analysis^11^. Moreover, the QNM hierarchical structure provides information on the relationships between multiple cell types *in vivo,* which is complementary to targeted spatial methodologies.

### Machine learning model to interpret age related changes in QNM topology

While robust data-driven network analyses can be used to reveal novel biology (unexpected co-expression of genes) in a manner agnostic to existing knowledge it is unclear if it provides greater insight into the molecular physiology of muscle compared with typical pathway-based interrogation of DE analysis. We describe above some of the features of the QNM built to represent human vastus lateralis muscle tissue from younger (n=270) or older samples (n=270) men and women. To generate a model that helps explain the influence of age on network structure (topology), we integrated both DE (e.g. DE after 2 weeks muscle unloading induced atrophy (UL47) or DE in primary skeletal muscle cells responding to 100nM Rapamycin) and QNM topological features from our five networks, excluding direct measures for gene scores in the young network. Rank order analysis indicated that DE values from short-term atrophy (UL47) show consistent global profiles with muscle frailty in 74yr people, but limited overlap with age driven DE (**Figure 3**). We developed a machine learning model using the topological relationships between DE transcriptional signatures, and network edges, including directionality, to establish if the key drivers for the topological re-wiring of older skeletal muscle were linked to the DE signature in a more sophisticate manner. In this context, topological re-wiring reflects age-related changes in how the transcriptome is coordinated within the muscle, capturing shifts in regulatory relationships that are not explained by or require expression changes.

#### Dramatic changes in network hub genes link to key physiology of aging

Manual inspection of the QNMs and DE signatures illustrated many distinct insights about muscle ageing not apparent with DE analysis. For example, *SZT2*, a subunit of the KICSTOR complex responsible for nutrient regulation^131^ went from the lowest 10% of hub gene connectivity values (strength) in young muscle (out of 605) to the top 5% in old muscle (out of 633). This dramatic shift makes it a novel candidate for mediating anabolic resistance (to nutrients) in aging muscle^132^. Critically, *SZT2* was *not* DE between young and old, illustrating the unique perspective QNMs provide. Another example is the Neddylation and aging linked gene *DCUN1D5* which dramatically *gains* connectivity in old muscle (**Supplementary Table 5**), while expression was *down-regulated* – and was down-regulated in both primary muscle and endothelial cells by Rapamycin treatment (see below). This implies that *DCUN1D5* re-wiring is pro-aging and subject to opposing compensatory down-regulation. Similarly, *BLTP3B* (also known as *SHIP164*) gained connectivity while abundance declined. *BLTP3B* is mutated in neurodegeneration, with a role in lipid transport and mitochondrial biology^133^. Given the scale of the data resource and models presented we built a machine learning model to help interrogate the age-related changes in network topology (**Table 1**, Y270 to O270) and how they link to muscle load networks (UL47, RT60, RT88), as well as key age (e.g. gene correlation with insulin resistance) and putative longevity phenotypes (DE responses to rapamycin treatment).

We built a machine learning model to predict the changes occurring within union of the top 600 ranked network genes in young (Y270) and older muscle (O270) networks (n=968 genes in total). The model used gene network strength measures, blinded from the Y270 network gene strength values (direct or indirectly), to predict change. Appropriate data scaling is critical, as topology metrics are not directly comparable across independent networks, so all cross-network topology and connectivity features are represented as within-network ranks or null-calibrated statistics^134^ (using null-calibrated proximity (NCP) and permutation based statistical modelling). Further care is required to interpret the meaning of the feature importance and model interpretability metrics (SHAP, SHapley Additive exPlanations values^135^). In Box 1 we provide a specific example of how the present analysis can be verbalised. In the present case SHAP analysis estimated each feature’s contribution (**Figure 8A**) to prediction of young to old gene rank topology changes (**Figure 8B**). We confirmed that most of the top features were weakly interrelated so that it would seem they provide independent sources of information to the model (**Figure S14A**). Aggregated at the block level (**Figure 8C and S14B**) we can see which data sources most strongly explain hub status transitions during aging. For example the intervention networks (unloading atrophy (UL47), hypertrophy responders in elderly (RT60) and hypertrophy responders in young (RT88)) as a group contribute most to the model (**Figure S14B**), while at the individual feature level some network level connections between DE genes (for genes down-regulated during long-term bed-rest and up-regulated by Rapamycin in cultured muscle cells) and the gene topology of the old network explain the rewiring of young to old muscle. Other highly ranked features include the positive correlating edges from insulin sensitivity genes and the rank order of gene strengths in the old muscle network and the importance of genes (by network strength) during atrophy and hypertrophy of muscle.

**BOX1.** *What is the connectivity calculation? For example, “DE_connectivity_Short_UL47” refers to a family of per-gene connectivity features that measure how strongly each gene is embedded, within a given topology network (e.g., O270, RT60, RT88, UL47), in the neighbourhood of genes that are differentially expressed in the Short_UL47 intervention. The Short_UL47 DE analysis defines two gene sets—genes that increase after unloading and genes that decrease after unloading (1.25FC and ∼1%FDR). For every gene in a topology network, we summarize its connections to these DE sets, separating positive and negative associations (reflecting the sign of the underlying edge weight). These connectivity summaries are converted into within-network ranks (the “_rank” values), so the model compares genes by their relative connectivity to the DE sets inside each network. The ranked features (e.g., pos_edges_to_Short_UL47_DE_up_O270_rank) are then grouped under the “Short_UL47 DE_connectivity” block for SHAP and ablation reporting. Separately, the “enriched module” idea applies to a different feature family (module_proximity / dist_to_enriched_module), where DE gene sets are used to identify enriched network modules (implying a specific biological function) and then compute proximity-type features.*

**Figure 8.**
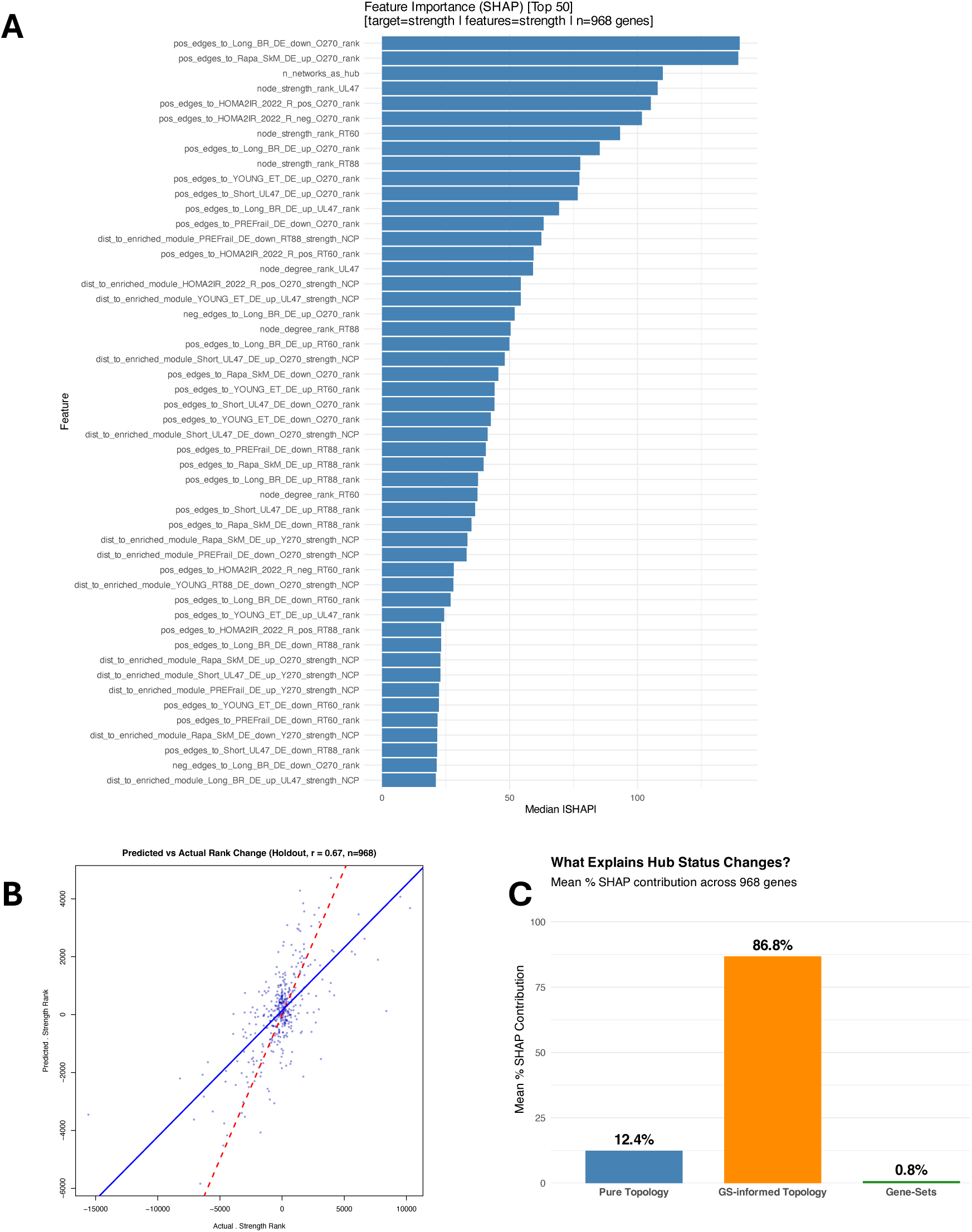
A) Plot of the top features that helped rank the network topology changes from young to old muscle (using the union of the top ∼600 in each network to build each model). B) Model spearman fit for predicting the change in gene network strength scores between young to old networks, in the absence of any information on the young gene rankings C) Contributions from DE gene set values, DE gene-set topology scores and the network topologies from 3 dynamic network models (atrophy, hypertrophy in young and old) to the model that helps explain the observed change in aging muscle topology.

We utilised ablation of features (**Figure S14C**), where a weak ablation score for an otherwise highly ranked feature, could indicate some unknown interaction: the model uses it, but removing it doesn’t hurt much because other sources of information substitute. In the present model, the Sex DE-connectivity and DE Insulin sensitivity (HOMA2IR^12^) contains information that reduces the prediction of Y→O topology rank change. In other words, genes that are highly connected (by ranked edge metrics) to the sex-DE gene set enriched modules are informative for which genes change topology rank from young to old implying that sex-linked transcriptional programs are aligned with the Y→O topology rewiring signal. It does not confirm that the Y→O transition is “sex-specific” because the block could be acting as a proxy for sex composition or other correlated structure in the dataset and overall, all the ablation effects were limited in impact.

The present model provides evidence that the biology of muscle atrophy and hypertrophy (especially in old muscle), along with the topological connection between insulin resistance and genes up-regulated by Rapamycin best define the rank changes in network wiring from young to old. It is possible to then examine the change in individual gene topology and which feature most influenced that change – and along with the network module contents and DE gene lists, then understand in detail how that gene responds to each age-related condition and which closely correlated partners may be influenced or regulating the observed changes. As can be seen in **Figure 9**, 124 genes with substantial shifts in topology, including some of the genes discussed in detail above (ARHGAP4, DCUN1D5, SDHB, SUCLG1 and SZT2) are now aligned to each group of features, so that the association with the machine learning interpretation of the data source can be directly observed. Interestingly, the topology changes from young to old, for the genes such as the mitochondrial related pair SDHB/ SUCLG1, have particularly strong links with our novel muscle pre-frailty signature – while the overall information gathered from the topological modelling of the long-term bed rest topology is self-evidence (Conn-LBR))

### Genome based and statistical models of muscle tissue ageing

So far, we have presented two main ways our muscle resource can be modelled to provide novel insight into human aging. Recently, it was reported that there was preferential targeting of long genes in aging^27,29^ and age-related disease^136^. Vermeij et al ^137^ proposed that longer genes may be more frequently down-regulated with age due to accumulation of DNA damage. DNA damage has also been proposed to be a stochastic driver^29^ for age correlated molecular changes, impacting on longer genes by way that they represent a greater absolute target – via events known as transcription-blocking lesions (TBLs). The main evidence comes from statistical modelling of RNA-seq data, which itself must be adjusted by a model that attempts to control for bias introduced by gene-length when using sequencing for detecting DE^138^.

Stoeger *et al* utilised RNA-seq to profile the transcriptome of 17 mouse tissues comparing 2yr with 4 months^27^. They acknowledged that methods used to adjust for length bias when using RNA-seq may limit the interpretation of associations with gene length. The median transcript length per gene was most consistently top ranked in their model, with the length of DE genes being most distinct between age 4 to 9 month (Spearman correlation of log_10_ transcript length versus age related fold-change). Stoeger *et al* reported that transcripts down-regulated with age were significantly longer in several mouse studies and in muscle from the GTEx dataset. Age related transcript imbalance was strongest in human frontal cortex, and weak for human muscle, reversed for human liver while 7 from 11 anti-aging interventions preferentially found increased expression of long genes (including Rapamycin and FGF21 treatment). In vivo partial reprogramming reversed the transcript imbalance but in the opposite direction from all other studies, with longer genes being enriched for increased not decreased DE by age. Olga Ibanex-Sole et al examined scRNA-seq mouse and human data to build support for this^29^. To establish an interaction with gene length and age-related DE, they used regression models and applied them to quartiles of gene-length; finding a significant association in all organs tests (very small n, and only males). They reanalysed the data from Peters *et al*^139^ – a blood age correlation signature (which has multiple disease influences) and reported modelling a list of 1,618 “down-regulated” genes and 1262 “up-regulated” genes. It is not clear what this analysis represents, as meta-analysis of discovery and replication data identified 897 negatively and 600 positively *correlating* genes with age, not DE.

These recent studies focused on protein coding genes and so it is also unclear if the same aging pattern is true for noncoding genes. Our resource can be used to model age related changes in gene expression without any confounding gene-length issues within the DE modelling. Our custom CDF processed high density array signal does not have a gene length bias problem (dedicated probes and no link between length and signal), while it also provides greater coverage of lncRNAs^6,11,12,32^ in muscle. We modelled the relationship between protein coding and lncRNA gene length and DE with age, as well as in response to Rapamycin treatment. The relationship found between protein coding gene-length and upregulated, down-regulated and unchanged gene expression between younger and older muscle tissue (n=540, **Figure 10A**) did not support the earlier analyses. In fact, we find the opposite result. The distribution of gene lengths (on a log10 scale) within each expression category (upregulated, downregulated, not significant) is presented as a probability density function). Protein coding genes up-regulated with age were longer (4.8kb log10) than genes down-regulated (4.46kb log10, 3×10^-19^ KS p-value). The pattern with lncRNA was more complex, where genes up-regulated with age were longer (4.66kb log10) than lncRNAs unaltered with age (4.17kb log10, 1×10^-7^ KS p-value) as were genes down-regulated with age (4.46kb log10, 2×10^-3^ KS p-value). We found no significant pattern with Rapamycin treatment. We also studied Pseudogenes, but these are heterogeneous in origin (e.g., processed vs. unprocessed) and structure where processed pseudogenes are shorter (because they’re retro-transposed and lack introns), while duplicated (non-processed) pseudogenes are longer and this led to multimodal plots and no obvious pattern.

**Figure 9.**
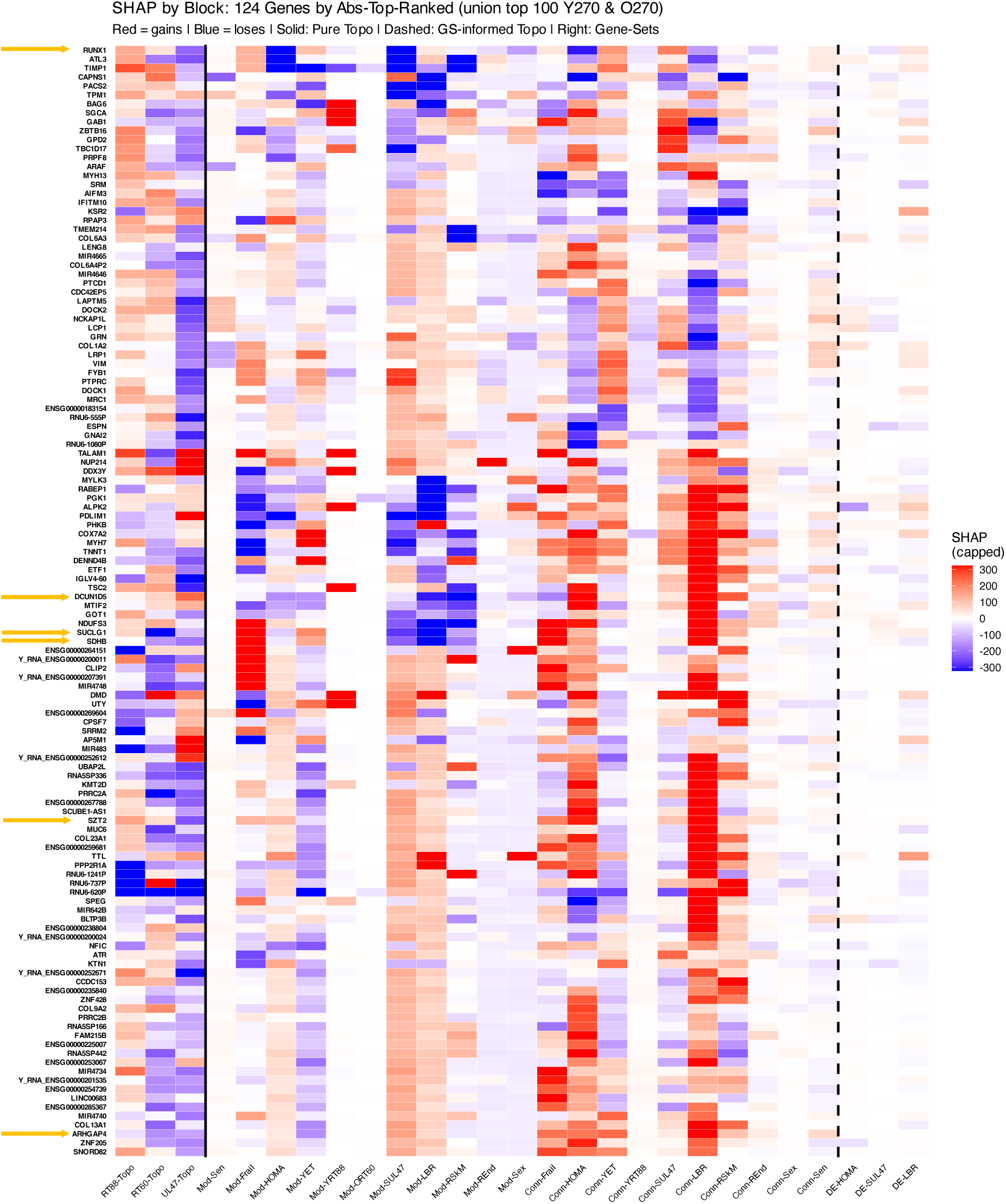
Examples of top changing genes with age, and their links to model input features by group. Yellow arrows highlight some genes discussed in the manuscript.

**Figure 10.**
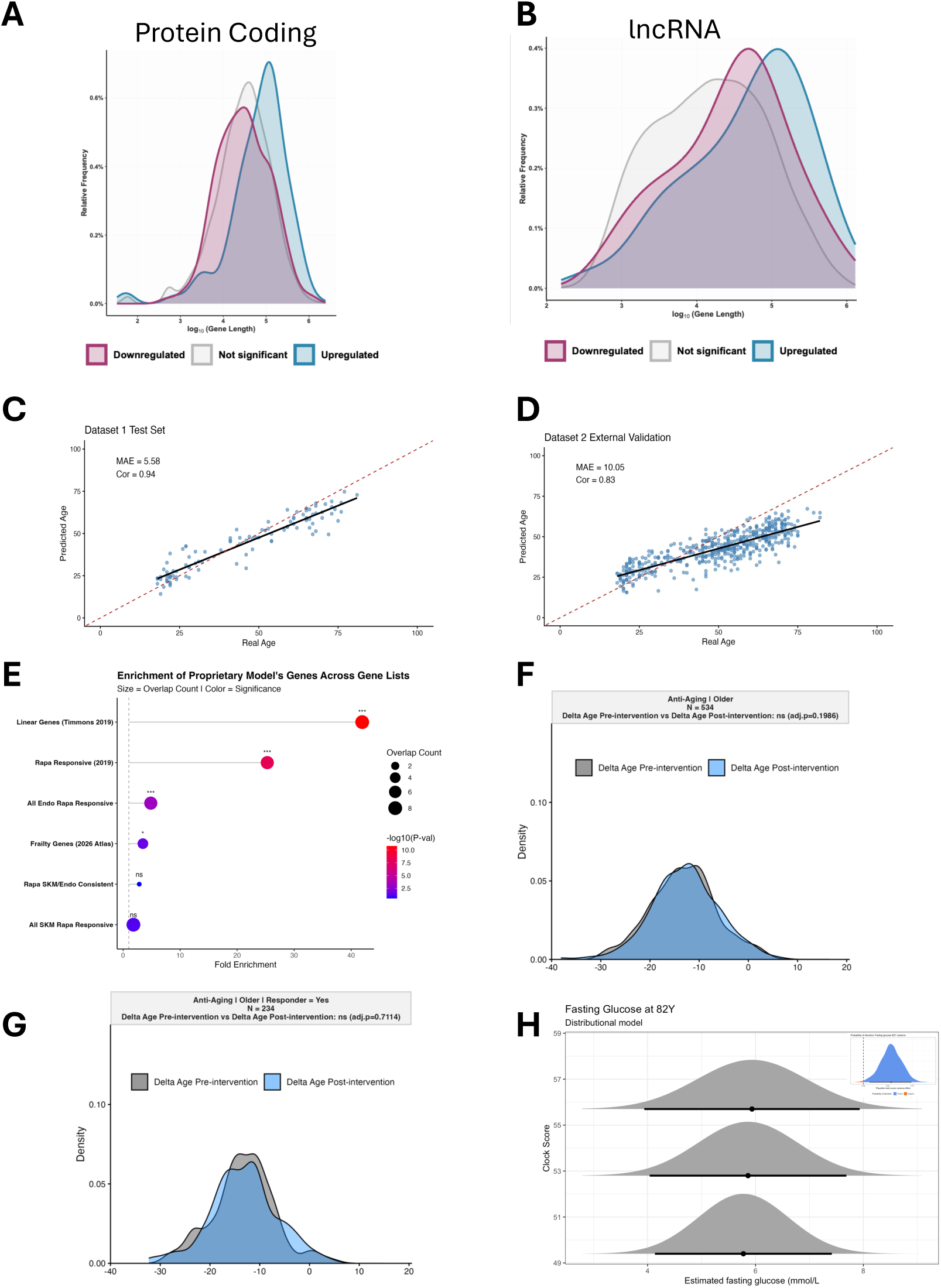
A and B) Protein coding genes up-regulated with age were longer (4.8kb log_10_) than genes down-regulated (4.46kb log_10_) 3×10^-19^ KS p-value. B) lncRNA up with age were longer (4.66kb log_10_) than lncRNAs unaltered with age (4.17kb log_10_) 1×10^-7^ KS p-value. lncRNA genes down-regulated with age (4.46kb log_10_) were also longer than lncRNAs unaltered by age (2×10^-3^KS p-value). Our data do not support the hypothesis that longer genes are more susceptible to damage induced loss during human ageing. B) A transcriptomic age clock was built using n257 biopsy samples and validated in independent pre-intervention samples (n=498) and applied to post-intervention samples (n=591) where post-intervention represented the biopsy after exercise (+ve) or unloading (-ve) interventions (with multiple studies have single-leg intervention paradigms). The model overlapped with a previous linear analysis of tissue age, and genes deemed rapamycin responsive. The model reported age systematically lower (p=0.0074 C 0.0044) than chronological age based on 10K boot-strapped aggregated age distributions and no intervention (positive or negative) altered ‘clock age’. Sub analysis also revealed no positive impact of RT on predict age nor a negative one from unloading. H) ULSAM 18y longitudinal study

Regression models, combining molecular features into an ensemble model, typically called a ‘clock’ has been one of the most active areas of mammalian research over the past decade. In some cases, the ‘clocks’ are repackaging of well-established epidemiological biomarkers for health and age-related disease; some are ensembles of DNA methylation^140^ or other molecular markers (sparse models, that evolved yearly), while the higher performing regression models tend to be recent combination of both^141^. Most of the recent analysis indicates that most clocks developed from 2015 until 2021 are considered defective (by the same people that published hundreds of articles using them)^142–144^. Indeed, it’s possible to skip the bisulfite conversion step when using the Illumina “DNA methylation” array and still yield robust chronological age clock - suggesting that this genotyping chip suffers from indirect influences^145^.

Recently, Ikram argued that ‘biological aging’ clocks in health research were being misused^146^ and that aging clocks should identify aging biological pathways (classification models should be sparse, to avoid over-fitting, and thus by default are not ideal for pathway over-enrichment analysis). Building an aging clock with a perfect fit with chronological age means it no longer offers an ‘error’ value that can be correlated with clinical status (as the measure of biological age). The driver for the “error” in a model is typically never established nor distinguished from confounding clinical variables. Recently it has been argued that biological age can be established from stochastic variation alone, Meyer’s and Schumacher argue that CpG epigenetic clocks measure how disordered a system has become^147,148^. The statistical validity of how most clocks have been applied to test for associations appears unsound^149^, and an urgent re-evaluation of all molecular age clocks validity is probably merited. This is illustrated by unstable outcomes achieved with some blood-based transcriptomic clocks, where a >3yr reduction in blood ‘biological age’ was achieved with only 4 weeks exercise, while control subject got >3yr older in the same 4 weeks^150^. Further, despite aggregating data from many independent labs, totalling >900 muscle profiles, Voisin et al ^151^ produced a muscle DNA methylation clock that was found not to be distinct from a random signature, when independently tested^152^. So how age epigenetic clocks can be made reliable over the longer term is not clear, and how “age clock” epigenetic modifications confer their influence on the biology of the cell remains uknown.

Interestingly, none of the epigenetic clocks reliably predict changes in the transcriptome – with often chance level observations being made in small subsets confounded by disease, from a larger multi-platform meta-analyses. For example, Voisin et al^153^ reported age-associated transcriptome analysis involving “>3,000 muscle samples”, that in the end modelled transcriptomic age data from 225 samples, half of which were compromised by metabolic disease or had incompatible genome models. Meyer and Schumacher have reported that epigenetic and transcriptomic age clocks can be built on the stochastic variation that occurs during aging^154,155^. We found no published^156^ based transcriptomic clock that worked with our human skeletal muscle data (the data we use is from a single technology platform with precise nucleotide level transcript alignment); so we built and validated our own tissue transcriptomic age “clock” and then evaluated its response, in the largest intervention data set to date, studying the influence of load status (exercise or unloading).

A transcriptomic age clock was built using n=257 biopsy samples independent of all other samples used (R=0.94, MAE=5.58, Seed=1010, alpha=0.5 and n-folds=10). We applied the identified 60-gene ensemble to a large independent cohort that represented the pre-intervention samples (n=498) i.e. the baseline samples from individuals subject to exercise or muscle unloading. We did not train on this larger data set as we wanted to retain the pre and post samples as a completely independent data source. We calculated ‘transcriptomic age’ in the post-intervention samples with the same 60-gene model (n=591; where post-intervention profiles represented a biopsy after exercise (+ve) or atrophy induced unloading (-ve) and multiple studies had single-leg intervention paradigms so there were two post-interventions samples for each baseline sample). The 60-gene model was able to significantly “predict” sample age (R=0.83), and interestingly it had a significant overlap with rapamycin responsive DE, and an overlap with an earlier linear correlative model adjusted from metabolic and cardiorespiratory fitness^11^ (**Figure 10C, 11D and 11E**).

Understandably, we evaluated older and younger subject separately for the impact of exercise training or muscle unloading and based on 10K boot-strapped aggregated age distributions, no intervention reliably (positive or negative) altered ‘clock age’ in people >50y (**Figure 10F**). Given we have proven that ‘responders’ to exercise have a distinct transcriptional response, we also did a sub analysis and again found no consistent impact of exercise (nor any influence subtypes of exercise separately, **Figure 10G**). Our transcriptomic clock was not interpretable in young people subject to muscle unloading or exercise, as both interventions – “good” and “bad” – resulted in a small *reduction* in clock age (**Figure S15A-C**). We next evaluated the clock’s relationship with cardiometabolic and musculoskeletal health in the ULSAM “70y” cohort^33^. Applying distributional regression and modelling mean and variance as a function of clock score we noted that 10yr following the muscle biopsy (and hence clock score) variability in glucose values were positively related to an older baseline clock score (after adjusting for small differences in chronological age, baseline fasting glucose and BMI, **Figure 10H**).

Thus, the present “transcriptomics age clock” was generally stable across >500 paired muscle samples (irrespective of physiological status). This suggests that our age-correlated rapamycin responsive transcript signature does indeed capture age correlated biology, as any “age-signature” modified by the vagaries of human exercise behaviours might not be a reliable linear correlate of chronological age. In short, it might be that a reliable clock ‘does not tick’ in the face of lots of short term ad hoc environmental influences. This thinking is in line with recent estimates, that indicate the influence of positive life-style, including exercise, on life-span probably equates to no more than 1yr^157^ and in general fits with realities of life span extension^158^.

## Summary Discussion

Taken together, by combining more comprehensive transcriptome coverage, with quantitative network modeling and integrating single-cell and spatial transcriptomic data sources, we provide arguably the most comprehensive, systems-level view of human skeletal muscle, adapting to load and age. We identified coordinated gene networks, cell-type–enriched transcriptional programs, and spatially organized molecular signatures that are not typically apparent from differential expression analysis. Together, these uncovered previously unrecognized hub genes that define human muscle, as well as introducing a mechanistic model of human skeletal muscle age. Instead of physically isolating cells for use with sequencing methodologies, we took several complementary approaches to identify cell specific events. First, we developed five quantitative network models representing ∼3 million cells obtained from muscle biopsies from men and women. Applying trillions of calculations, we identified the hub and module structures under multiple load conditions, discovering dominant hub genes previously unknown to skeletal muscle biology. These models integrated scRNAseq data and profiles of cultured single cell-types, to model cell-type enrichment across thousands of co-expression modules. We complemented this by using multiple spatial transcriptomics technologies, directly capturing the anatomical location of gene expression^159^ work that continues, with the aim of building spatial based models of muscle age and load. We identified a novel human age-related frailty signature, while network modelling revealed accurate senescence processes previously opaque in vivo^11,33^. Thus, our network approach reveals senescence processes, complements the scRNAseq based studies, illustrating single gene-gene interactions.

The machine learning model identified drivers of age-related network re-wiring, including interactions with insulin-action and Rapamycin. We illustrate the utility of this new human muscle resource, identifying novel relationships between gene length^27,28^, aging^27,29,30^ and protein coding status, while present a new transcriptomic age “clock” and its response to altered load status in >500 people. A major conclusion from the present work is the QNM level analysis and threshold free rank-based methods reveal more insight into skeletal aging biology, and how it relates to various factors that alter the physiological capacity of muscle, than traditional DE analysis (where reliability is heavily dependent on sample size and size-effect of the perturbation). We find that a major influence on muscle age remains muscle disuse which may reflect degeneration of the neuro-muscular interface, behavioural related reductions in muscle loading or both. We also discover that most of the most strongly inter-connected genes in human muscle have never been studied before in muscle, illustrating the power of network modelling to reveal entirely novel biology – including hub genes that re-wire during aging, under the influence of the established signalling processes. Finally, we identify that the paracrine acting secreted protein GDNF is robustly induced in human muscle during *disuse*, and this dissipates over time, while the ubiquitously measured myokine, IL6, is unlikely to originate from muscle *in vivo*. Release of the raw data, network data files and browsable versions of the modules, represents a powerful resource to facilitate new understand of human skeletal muscle aging.

## Methods

The network models relied on 930 profiles, the transcriptomic clock on 1,346, the longitudinal work on 105, the Rapamycin on 28 samples, including our previously released primary muscle data^11,160^ while 97% of the samples utilised came from our laboratories and <1% were biological replicates run at distinct times (**Supplementary Table 10**). Depending on the purpose of the analysis or whether it was paired or unpaired, a distinct Combat based batch correction was applied – reflecting the fact that there is not a one-size fits-all approach to data scaling and normalisation. For processing the data for the transcriptomic clock models, age was set as a protected factor, and the influence of study and sex was adjusted for. A total of 1,675 samples were utilised. It must be emphasised that in all studies, 1-2% of samples were removed after inspecting the PCA plots and during batch correction, but prior to any primary statistical analysis.

### Transcriptomic Methods

#### RNA processing

Typically RNA was extracted from 20 mg of muscle tissue^161–164^ in 1000 mL of TRIzol, using ceramic microbeads (MP Biomedicals, Solon, OH, USA) and homogenised using a FastPrep tissue homogeniser (MP Biomedicals, Solon, OH, USA) or Qiagen Tissuelyser. This was followed by chloroform extraction, and the upper aqueous phase transferred to an RNase-free tube. RNA was processed for transcriptome profiling using the GeneChip WT Plus Reagent Kit (no PCR). First and second-strand cDNA synthesis was performed using ∼100 ng of RNA and cRNA purified using magnetic beads. 15 mg of cRNA was then amplified and hydrolysed using RNase H (leaving single-stranded cDNA) and 5.5 mg was then fragmented, labelled, and hybridised to the HTA 2.0 platform. This was washed and stained using a GeneChip Fluidics Station 450 and scanned using a GeneChip Scanner 3000 7G.

#### Computational processing of the high-density exon-based platforms

We previously described a process for enhancing the processing of multiple transcriptomic technologies^164–168^. For example, the HTA 2.0 platform has 6.9 million 25-mer probes. The sequence of each probe is checked against the recent reference genome and transcriptomes^169,170^. To do this; we produce FASTA type (with a unique label for each 25mer probe), a probe GC content and a probe-level chip definition (CDF) files. The FASTA file was aligned against Grch38 - Gencode 43^171^ using the STAR aligner^172^. Probes that uniquely map were combined to form groups of probes (probe-sets). To assess probe signal performance, signal was scanned in >1000 individual profiles using a probe-level CDF (aroma.affymetrix^169^, affy^173^ and affxparser packages). Probes with both a very low signal and a low coefficient of variation were removed^164–168^. This optimised CDF was used to summarise RNA expression from a GC-corrected CEL file and iterative rank-order normalisation (IRON)^174^ in the default mode. From the original 6.9M probes, reannotation and summarisation yielded a transcript level probe-set CDF that used 5.23M probes to define 220.6K probe-sets prior to study-specific transcript signal filtering (each probe-set relied on n>3 probes; median = 66). Note that the public CDFs integrate probes with “dead” signals and we identify and remove those from contributing to the probe-set signal^12,31,32^ so enhancing the signal for thousands of transcripts, particularly are the lower end of the signal range (Figure S1A). Our custom CDFs are deposited along with raw data files and QC and batch corrected expression matrices (ArrayExpress). This updated CDF can be utilised to extract any other human muscle data sets produced on the HTA 2.0 platform. We estimated that a total of ∼165K transcripts were annotated above the background signal using standard deviation-based filtering. This did not include IL6 but did include satellite-cell expressed BDNF^110^ and the IL6 Receptor. We used SAMR^175^ with 10K permutations to estimate the false discovery rate (FDR), which is more robust than other FDR methods^176^. Paired or unpaird SAMR analyses was applied where appropriate. We typically relied on an FDR cut-off of 1% and a ≥ |1.25| fold change lists.

### Network building and Machine Learning interpretation Methods

Network Construction and Topology Feature Extraction: Gene co-expression networks were constructed using Multiscale Embedded Gene Co-expression Network Analysis (MEGENA), which employs planar filtered network construction and multiscale clustering to identify hierarchical modular structures (Song and Zhang, 2015). Separate networks were built for young (Y270) and old (O270) muscle tissue samples, as well as intervention networks representing unloading-induced atrophy (UL47), resistance training-induced hypertrophy in young muscle (RT88), and resistance training-induced rehabilitation in aged muscle (RT60). We also inspected a previous bulk muscle database – MyoMiner. Unlike Metamex, a robust gene alignment approach was taken, using a single 3’ RNA genomic platform, assigning the probes to gene (ENSG) level ensembl annotations (version 20.0.0) using a publicly available CDF. We find when checking some of our candidate genes, that >6,000 genes had a *significant* correlations, indicating there must be some sort of structural bias in that dataset.

A major overhaul of the plotting functions available for MEGENA was developed, focused on project the network structure on to a sunburst plot, developing an integrated heuristic for combining scRNAseq data ^49^ and number of bulk-method profiled single cell types (HTA 2.0 platform)^31^, the bulk signal from muscle (thresholding so called cell type specific marker genes, that should be low expressed but were in fact abundant in the bulk signal) to yield a cell type enrichment model applied to each module. Additional statistical enrichment for hypothesis driven gene-signatures was applied, to characterise modules, and in the case of the machine-learning model, define distances between important genes e.g. hub genes, and enriched modules – across networks, by edge and directionality (calculated separately from MEGENA, where Rho is not given a direction).

For each network, MEGENA computes node-level topological properties including strength (sum of edge weights) and degree (number of connections). In addition to the network models themselves, there were additional in vitro and in vivo data sources (e.g. Rapamycin treatment or insulin resistance correlated genes) and in total 22 “phenotypes” were processed and this yielded >700 phenotypes (**Supplementary Table 11**). Many of the network characteristics cannot be directly compared across networks (due to differences in scale) and so rank and/or statistical representations (of distances) were required to contrast topology metrics, so all cross-network topology and connectivity features are represented as within-network ranks or null-calibrated statistics^134^ (using null-calibrated proximity (NCP) and permutation based statistical modelling). Further care is required to interpret the meaning of the feature importance and model interpretability metrics (SHAP, SHapley Additive exPlanations values^135^) and we utilised heatmaps and feature ablation to improve the interpretability of the analysis. In the model code there’s an explicit rank-data validation step that checks all numeric columns beginning with distance topology labels show a maxima roughly bounded by the number of genes, i.e. behave like rank variables to prevent absolute-value metrics being used. An example of the approach to verbalise a result would be: “*Connectivity during muscle unloading to modules enriched in Rapamycin differentially expressed genes (ranked within network) substantially contributes to the modelling Y→O topology rank change (assuming that this feature is not strongly correlated with other features – see heatmap) suggesting that the biology captured by this relationship may help explain age-associated network rewiring*.”

Objective: The model was trained as an XGBoost regression, on the numeric rank deltas (rank_Y270 − rank_O270) for muscle age. This ranked the genes that undergo significant changes in connectivity rank between the Y270 (young) and O270 (old) networks so that we can attempt to explain the young-to-old network re-wiring. The target is rank change (rank_Y270 minus rank_O270) rather than absolute topology delta, as rank changes capture biologically meaningful transitions in hub status that can be compared across networks. Training and prediction were restricted to genes in the union of those within the top 650 by strength in either the Y270 or O270 network (union of the two top-650 lists), yielding *n* genes in the training set. This choice balances including a broad set of hub and near-hub genes while keeping the model focused on genes with high network connectivity, thereby capturing stable hubs (top in both networks), genes losing hub status (top in Y270 only), and genes gaining hub status (top in O270 only). Positive target values indicate higher connectivity rank in young (Y270) and negative values indicate higher rank in old (O270).

Feature Engineering: Features were also organized into interpretable blocks, with strict exclusion of Y270 values to prevent information leakage. Two independent toggles controlled feature selection: “TARGET_TYPE” determines which rank delta to predict (strength or degree-based topology values), and “FEATURE_TYPE” determines whether strength or weight features are used by the model (this is to reduce redundancy since strength and weights are derived from the same edge data and partly related). Leakage prevention and feature exclusions included:

1. All features derived from the young network (_Y270), including absolute values and any young-versus-* deltas (_Y270v*).
2. Any direct young-versus-old deltas (_Y270vO270), which can trivially reconstruct the target.
3. Absolute topology values from the old network (O270) for the same measure as the target (to avoid tautological predictors).
4. Differential expression results generated from the same samples used to build the Y270/O270 networks (e.g., AGE540_DE), to avoid sample overlap leakage.

Topology blocks: These comprised network topology features (absolute topology in intervention networks RT88, RT60, UL47; O270 vs intervention deltas O270vRT60, O270vRT88, O270vUL47 comparing old muscle to each intervention condition. This relyied on the conserved hub connectivity for intervention networks) and Gene Set-informed Topology features (module proximity distances) using Null-calibrated proximity and permutation-based statistics, to phenotype-enriched modules.

Differential expression blocks: Direct DE values from phenotype studies (e.g. pre-frailty, insulin sensitivity (correlation), hypertrophy in young or old people, gains in VO2max in adults, short-term unloading (1-2wks, with older people 1wk, and younger people 2wks) induced atrophy or long-term bed rest (younger healthy subjects, 84 days), rapamycin responses in cultured muscle or endothelial cells, DE for sex and literature curated senescence markers. Connectivity to DE gene sets (edges and weights to upregulated/downregulated gene lists); and changes in DE connectivity were calculated. The O270 vs intervention deltas have specific biological interpretations: e.g. O270vRT60 captures how old muscle differs from rehabilitated old muscle (“exercise reversal”), O270vUL47 captures how old muscle differs from unloaded muscle (“disuse”), and O270vRT88 captures how old muscle differs from young hypertrophying muscle.

Model Training: Gradient-boosted decision trees^177^ were trained with a stratified 60/40 train-holdout split based on joint quantiles of Y270 strength rank and degree rank. Hyperparameters were tuned via 5-fold cross-validation optimizing Spearman correlation between predicted and actual absolute rank changes, with early stopping to prevent overfitting. Parallel processing was used for hyperparameter grid search, and bootstrap stability analysis. Hyperparameters were selected to maximize Spearman correlation of absolute rank change (|predicted| vs |observed|) during cross-validation, and performance was reported on a held-out test split.

Feature importance was assessed at the block level through two complementary approaches:

1. Block ablation: For each feature block, the model was retrained with that block removed and the drop in holdout Spearman correlation was measured, normalized by feature count for fair comparison. This provides causal evidence of each block’s contribution to predicting rank changes. Additive reconstruction: Starting from no features, blocks were progressively added in order of per-feature ablation importance. This reveals the cumulative prediction accuracy with topology blocks alone versus with all blocks, quantifying the marginal contribution of differential expression features. Gene-level interpretation was achieved by partitioning each gene’s SHAP values into topology and DE contributions, yielding topology_pct and DE_pct for each gene
2. Robustness Checks: Bootstrap stability was assessed by retraining on 100 bootstrap samples and identifying features appearing in the top 20 most important in at least 70% of runs. Permutation importance was calculated on the holdout set by scrambling each feature and measuring the increase in RMSE.

Interpretation: Feature importance and model interpretability were assessed using SHAP (SHapley Additive exPlanations) values^135^. SHAP analysis quantifies each feature’s contribution to individual predictions, aggregated at the block level to identify which feature families most strongly explain hub status transitions during aging. The final output includes gene rankings with explanation partitioning: each gene receives a predicted rank change score, along with the percentage of that prediction attributable to topology versus differential expression features. GO enrichment analysis was performed to identify biological categories preferentially explained by topology or DE features based on median explanation percentages.

### Spatial Methods

We utilised three distinct spatial technologies^72,74,77^, as described above. This allowed us to generate sequencing libraries from 57 areas of interest, across four slides (three replicate slides of eight baseline samples from Stokes et al^24^ and one slide with eight post-unloaded muscle (7 of which were intact). We profiled eight samples across two slides (pre- and post-unloading) using Xenium and profiled 60 biopsy samples (ROIs, pre- and post-unloading) using the MERSCOPE technology and V1 chemistry, of which 54 ROIs were intact. The samples originated from the unloading studies described below.

We used two spatially resolved single-cell transcriptome profiling technologies – Merscope and Xenium. As part of the process of designing and validating ongoing work in our laboratory, we produced images of the location of a small number of the ncRNAs identified in the present study. The MERSCOPE assay also included markers for muscle fibre types, (type I fibres [MYH7 fish probe]^178–180^, type II fibres [ATP2A1 fish probe]^179,180^, satellite cells [PAX7, MYF5 merfish probes], and endothelial cells [ENG, TIE1, PECAM1, APLNR merfish probes])^178,181^. The panel was used to profile three independent muscle samples originating from the present study, and some ncRNA data pertinent to the present results is presented. The gene list was sent to Vizgen Inc, and they selected 30-50 merfish probes per gene for profiling on the MERSCOPE. Briefly, each fresh frozen sample embedded in the optimal cutting temperature compound (OCT, Tissue-Tek, The Netherlands) was sectioned at a thickness of 10 μm in a cryostat at -20 °C and transferred onto the circular MERSCOPE glass slide (PN 2040001, Vizgen, USA) within the fiducial bead border, and allowed to adhere to the glass for 5 minutes^77,182^. Tissue sections were fixed in 4% paraformaldehyde (PFA) at room temperature for 15 minutes. After fixation, the tissue sections were washed 3 times in 1X PBS for 5 min at room temperature, and permeabilized in 70% ethanol (molecular grade) at 4°C overnight. Subsequently, the tissue autofluorescence was quenched in the MERSCOPE Photobleacher (PN 10100003) for 3 hours at room temperature. Then, the 70% ethanol was removed, and the sections were washed with 5 ml 1X PBS and subsequently stained with Vizgen cell boundary staining kit (PN 10400009, Vizgen, USA). Briefly, sections were incubated in Blocking Solution (PN 20300100, Vizgen, USA) and cell membranes were stained with Cell Boundary Primary Stain Mix (PN 20300010). After 3 washes in 1X PBS, Cell Boundary Secondary Stain Mix (PN 20300011) was applied to the tissue sections. RNase inhibitor was added to each step, and all incubations were performed for 1 hour at room temperature. Afterwards, the tissue sections were fixed with 4% PFA for 15 minutes at room temperature. After 2 washes in 1X PBS, the tissue sections were hybridized with the custom-designed merfish panel at 37°C for about 40 hours. Then, the slides were incubated twice in the Formamide Wash Buffer (PN 20300002) at 47°C for 30 minutes and rinsed with the Sample Prep Wash Buffer (PN 20300001). Subsequently, tissue sections were embedded in polyacrylamide gel to immobilize RNA and cleared in clearing premix (PN 20300003) containing Proteinase K for 24 hours at 37 ^0^C until the tissue sections became transparent. Finally, the tissue sections were stained with the DAPI and PolyT Staining reagents (PN 20300021) for 15 minutes at room temperature. The hybridized probes were imaged using the MERSCOPE (Vizgen Core Laboratory, Boston, USA) with default settings (PolyT and DAPI and Cell Boundary Channels “on”, while scan thickness was 10μm). The raw images were converted to “vzg” meta-output files. For ncRNAs included in both assays, we used the MERSCOPE Vizualizer software and cell-specific marker probes to evaluate the location of each RNA. The Xenium profiling was arranged by 10X Genomics and processed at the University of Oxford, using the standard Xenium protocols and a custom 460-gene panel designed from our 960-plex Merscope assay. GeoMX data was generated at the Seattle HQ of Nanostrings as described by Merritt et al^74,183^ with the muscle tissue sectioned (10um) at -20, and stained using three antibodies (Nuclei (SYTO13), Type II fibres (MYH2) and Endothelial cells (CD31)). Raw sequencing reads collected from each AOI, were pooled and sequenced, and % Aligned, % Trimmed, or % Stitched Sequencing Read thresholds applied as per the DSP workflow^74,183^ (this protocol yields ∼80% PCR duplicates).

### Gene Length and Aging Clock modelling

The relationship between protein coding gene-length and upregulated, down-regulated and unchanged gene expression between younger and older muscle tissue (n=540, **Figure 10A and 10B**) does not support the earlier analyses. The distribution of gene lengths (on a log10 scale) within each expression category (upregulated, downregulated, not significant) as a probability density function. The relative frequency density — the integral over the curve equals 1 (so the Y-axis is relative frequency per log10(bp)). Each line shows the distribution (not count) of gene lengths within each category, scaled so the area under each curve is 1. KS test is non-parametric and compares the entire distribution (good for shape and shift). Wilcoxon rank-sum (Mann– Whitney) tests medians (good if distributions are similar but shifted). For spearman plot inverse hyperbolic sine was applied to compress the FC values at extremes. The transcriptomic aging “Clock” is an ensemble regression model built using R’s glmnet package (v4.1-10). We applied 10-fold cross validation to automatically select the best lambda of 3.06573 using mean squared error. The alpha was set to 0.5 and the seed was set to 1010 for reproducibility. With an L1 ratio of 0.5, we applied the elastic net model to 60% of the training dataset to select the best lambda, and further validated it on the remaining 40%. We utilised the “clock” to evaluate if either muscle loading (exercise) or unloading (atrophy/disuse) nudged the distribution of clock scores. We also evaluated if the “clock” was related to long term metabolic health using both a Bayesian and Frequentist methodology. We used Bayesian distributional modelling^184^ with the brms R package^185^ to examine the effect of clock score at ∼70Y on fasting blood glucose at 82Y and 88Y. The model for both mean and variance included age as a covariateas recommended by Krieger et al.^186^ and fasting blood glucose and BMI at 70Y to control for baseline influences^187^. Default priors were including flat priors on model coefficients were used. We also used distributional modelling^188,189^ via the glmmTMB R package^189^ to examine the effect of clock score from the biopsy obtained at ∼70Y, on fasting blood glucose at 82Y and 88Y. The model for both mean and variance included age as a covariate and fasting blood glucose and BMI at 70Y as above.

### Cross-sectional and intervention clinical cohorts

All studies were approved by their respective local research ethics boards and adhered to the ethical standards for research involving human participants as outlined in the Declaration of Helsinki. All participants provided both verbal and signed written informed consent. Several independent clinical studies contributed to the development of the present muscle transcriptomic resource and the original clinical publications should be referred too and cited. Here we present a brief overview of the studies. Four major exercise intervention studies provide the backbone to the current analyses.

#### There was one multi-centre META-PREDICT cohort

The study was funded by an FP7 grant written by Timmons and the trial was approved by local ethics committees at each centre (the University of Nottingham Medical School Ethics Committee: D8122011 BMS; the Regional Ethical Review Board Stockholm: 2012/753-31/2; the ethics committee of the municipality of Copenhagen and Frederiksberg in Denmark: H-3-2012-024; Comite etico de Investigacion Humana de la ULPGC: CEIH-2012-02; and the Loughborough University Ethics Approvals Human Participants Sub-Committee: 12/EM/0223). The MP cohort consisted of 189 *active* participants, recruited as previously described from 5 geographical regions across Europe^190^ working with standard operating procedures (SOPs). Subjects were (<600 metabolic equivalents (METs) min·wk^-1^)^191^ and had a fasting blood glucose level consistent with World Health Organisation (WHO) criteria for impaired fasting glucose (6.1 - 6.9 mmol·l^-1^), and/or a BMI >27 kg·m^-2^ but were free of clinical diagnosis or drug treatment for active cardiovascular or musculoskeletal disorders. HIT exercise training was supervised and consisted of three high intensity cycling sessions (5-by-1 minute at 125% VO_2_ max) per week for 6 weeks^190^. Prior to the baseline assessments, participants were instructed to refrain from exercise for three days.

#### There were three STRRIDE Cohorts

STRRIDE I and AT/RT study protocols were approved by the institutional review boards at Duke University and East Carolina University. The STRRIDE-PD study protocol was approved by the institutional review board at Duke University. The STRRIDE AT/RT (S2) cohort (NCT00275145) examined the effects of aerobic and/or resistance exercise on cardiometabolic health in sedentary, overweight adults randomized to one of three training groups for eight months: 1) aerobic training at 65-80% VO_2peak_; 2) resistance training 3 days/week,

8 exercises, 3 sets/exercise, 8-12 repetitions/set; and a combination of the aerobic and resistance training programs^41^. For the purposes of the present analysis and given the striking influence of responder status on the transcriptomic response^7,9^, subjects were categorised by their phenotypic outcome rather than their training protocol (which was not an obvious influence). The STRRIDE PD (S-PD) cohort (NCT00962962) involved older sedentary overweight adults without or without impaired fasting glucose but no medical treatment for active disease. The intervention groups included a) low amount/moderate intensity (10 KKW at 40-55% VO_2reserve_), b) high amount/moderate intensity (16 KKW at 40-55% V̇O_2reserve_); c) high amount/vigorous intensity (16 KKW at 65-80% VO_2reserve_) and d) clinical lifestyle prescription, which included low amount/moderate intensity exercise of 10 KKW at 40-55% VO_2reserve_ plus diet to achieve 7% body weight loss (comparable to the Diabetes Prevention Program (DPP)^40^. Again, for the present analysis either baseline data was used or in the case of the transcriptomic age clock analyses, responder status for either VO2max gains or hypertrophy were utilised.

#### São Paulo Elderly resistance training study

(Roschel is the contact for clinical information) represent muscle biopsies were obtained from the study by Lixandrao et al^192^. A total of 83 healthy older adults completed 10 weeks of unilateral resistance training (RT), which consisted of two supervised leg extension sessions per week. One leg performed a single set, while the contralateral leg performed 4 sets of 8 to 15 repetition maximum (RM). The present study focused exclusively on the leg performed 4 sets for DE analysis, as this condition represented adequate training and led to greater average muscle hypertrophy^192^ while all paired samples were considered in the age clock analyses. Of the 83 participants, paired biopsy samples before and after training were available from 60 participants (30 males, 30 females; 69 ± 5 years old; BMI of 26 ± 4 kg/m^2^). Participants consumed a whey protein supplement throughout the study – 20g of the protein twice daily, once in the morning and once 30 minutes before bedtime to ensure adequate nutrition in these older people. To evaluate hypertrophic responsiveness to RT, quadriceps cross-sectional area (CSA) was measured using magnetic resonance image (MRI) before and after the 10 weeks of RT. Skeletal muscle biopsies from the *vastus lateralis* were collected both pre- and post-training, following the MRI scans.

#### There were four clinical muscle unloading studies used

(Stokes, Mcleod, Phillips and Lim are the contacts for clinical information). Muscle biopsy tissues from these five independent clinical studies (two of which the omic data has been published^193–195^) were available, totalling 116 pre- and post-unloading samples from 58 young and old adults: 47 young men (24 ± 5 years old), 11 older men (67 ± 3 years old). All muscle biopsy tissues were obtained from the *vastus lateralis* using a 5-mm Bergström needle custom-modified for manual sectioning. For each participant in the present study, changes in thigh lean mass (LM) between pre- and post-unloading were assessed using dual-energy X-ray absorptiometry (DXA) to define unloading-induced muscle loss. Figure 1 shows a schematic overview of each study protocol. Despite differences in the duration of unloading (2 weeks or 84 days for young men and 1 week for older men) and in the presence of co-interventions such as contralateral leg exercise or nutritional supplementation, the relative unloading-induced reduction in thigh LM was consistent across studies, ranging from 5.0 to 18.1%. In the present study, RNA was extracted from all 116 muscle biopsy tissues (58 matched pre-post pairs) for transcriptome analysis using the HTA 2.0 array analysis.

For one study, muscle biopsies originated from Lim et al^193^ where nutritional changes were unrelated to atrophy outcomes. Active young men wore a knee brace (X-ACT ROM knee brace, Don Joy, Dallas, TX, USA) on a randomly selected leg for 2 weeks, constituting the unloading period. During this single-leg unloading phase, participants consumed standardized meals that consisted of 1.2g of protein/body mass/day, and crutches were provided to allow for mobility. Muscle biopsy tissues were available at pre- and post-unloading from 18 participants (21 ± 1 years old; BMI of 24.3 ± 2.9 kg/m^2^), including 10 in the Fortetropin group and 8 in placebo group. The Fortetropin group ingested 19.8 g of Fortetropin – a freeze-dried, fertilized egg yolk product – daily with breakfast, while the placebo group consumed cheese powder matched for energy and macronutrients. Thigh LM was assessed by DXA at pre- and post-unloading. The loss of thigh LM induced by single-leg unloading did not differ between groups; no effect on changes in muscle mass from the nutrition supplementations. A second set of muscle biopsies originated from Stokes et al^194^. Recreationally active young men performed unilateral leg extension and leg press exercises (3 sets of 8-12 repetitions, 3 days/week) on a randomly assigned leg for 10 weeks. During the final 2 weeks of the study, a knee brace (X-ACT ROM knee brace, Don Joy, Dallas, TX, USA) was applied to the contralateral leg – the single-leg unloading period. Participants consumed an average of 1.6 ± 0.9 g/kg/d of protein, including 25 g of whey protein isolate during the unloading period. Crutches were provided to allow for mobility. Muscle biopsy tissues were available at pre- and post-unloading from the unloading leg in 12 participants (20 ± 3 years old; BMI of 23.8 ± 3.1 kg/m^2^). Thigh LM was measured using DXA at pre- and post-unloading. There was no cross-talk or prevention of unloading-induced muscle loss from the contralateral leg exercise training ^194^. A third study used the same approach as Stokes et al^24^ but has not been published in any format. Six healthy young men (20 ± 1 years old; BMI of 24.6 ± 2.6 kg/m^2^) were recruited. The participants wore a knee brace (X-ACT ROM knee brace, Don Joy, Dallas, TX, USA) on a randomly selected leg for 2 weeks, constituting the unloading period. Meanwhile, the contralateral (non-unloaded) leg performed 4 sessions of unilateral leg extension and leg press exercise (4 sets of 8-12 repetitions), spaced over the last 8 days of the unloading period, with 24 hours of rest between each session. During the 2 weeks of unloading period, participants consumed a standardized diet containing 1.0 g of protein/kg/d. Crutches were provided to support mobility. Muscle biopsy tissues were collected pre- and post-unloading from the unloading leg, and the thigh LM was assessed using DXA scan. As confirmed in our previous study ^194^, acute exercise of the contralateral leg did not induce cross-talk or prevent unloading-induced muscle loss. A fourth study came from a second unpublished cohort (UL_OIM_HTA) where 11 healthy older adults (9 Males, 2 Females; 67 ± 3 years old; BMI of 26.7 ± 3.1 kg/m^2^) were recruited. The participants wore a knee brace (X-ACT ROM knee brace, Don Joy, Dallas, TX, USA) on a randomly selected leg for 1 week, constituting the unloading period. During this single-leg unloading phase, participants consumed standardized meals that consisted of 1.0 g of protein/body mass/day, and crutches were provided to allow for mobility. Muscle biopsy tissues were available at pre- and post-unloading. Thigh LM was assessed by DXA at pre- and post-unloading. We did not use the change in lean mass values due to the uncertainly to measuring changes over only 1-2 weeks with the DXA methods employed in these studies.

A fifth disuse study was the reprocessing of an 84-day bed-rest study (UL_BR-84days). The transcriptomics arrays were funded by our META-PREDICT study. Muscle biopsies came from the study by Fernandez-Gonzalo et al^195^. Here, healthy young men underwent 84 days of bed rest without any exercise (or limited exercise, which we did not utilise). Participants lay in the 6° head-down tilt position. Muscle biopsy samples were obtained from the vastus lateralis of the dominant leg at pre- and post-84-days of bed rest in 11 participants (32 ± 3 years old; BMI of 23.3 ± 1.9 kg/m^2^). The original transcriptomics analysis was inadequately processed by Rullman et al^195^, using a suboptimal QC process and CDF – and as a result we identified 500% more DE than the original study reports, including identifying some of the largest gene expression response to unloading, missing from the original article.

#### Muscle Pre-Frailty

A transcriptomic analysis of older adult muscle with or without evidence of developing frailty was originally published in a study led by van Loon^98^. We published this analysis of muscle frailty using a low-sensitivity array^196–200^ which we have replaced with the HTA 2.0 platform and novel pipeline. Frailty status was defined using the Fried criteria^201^, which identifies individuals as frail if three or more of the following conditions are present: unintentional weight loss (>=10 lbs in the past year), poor endurance and energy (self-reported exhaustion based on CES-D scale questions ^202^), weakness (grip strength in the lowest 20% at baseline), slow walking speed (lowest 20% based on a 15 feet walk test), and low physical activity (lowest quintile of a weighted score of kilocalories expended per week, calculated based on self-reported physical activity ^203^). The new transcriptomes were obtained from 32 frail older adults (17 males, 15 females; 73 ± 6 years old; BMI of 28 ± 4 kg/m^2^; leg press 1RM of 136 ± 32 kg; 49 ± 9 kg of total muscle mass) and 36 healthy older adults (19 males, 17 females; 71 ± 5 years old; BMI of 26 ± 3 kg/m^2^; leg press 1RM of 179 ± 42 kg; 53 ± 12 kg of total muscle mass). These represent baseline muscle tissues and the unpaired SAMR contrast generated a frailty-associated gene signature that was more robust than the earlier analysis and we found to strongly relate to short and longer-term muscle disuse (**Figure 3 and Figure S5**).

#### Rapamycin (100nM) studies (Primary human myotubes and HUVEC cells)

The primary muscle cells were produced and profiled in our lab and the methods are reported elsewhere^11,24,160,204^. The transcriptomic data was updated to the same references using the CDF described above. HUVECs (Promocell Cat#C-12203; pooled donors) were seeded into 6-well plates and were grown to >95% confluency (37°C; 5% CO_2_). Once confluent, media was removed and replaced with Endothelial cell basal medium (EBM) (Promocell; C-22110) containing 2%(v/v) fetal bovine serum (FBS) and 100nM Rapamycin for 24 hours with EBM + 2% FBS serving as control. After 24 hours, media was removed, and cells were washed with 1X phosphate buffered saline (PBS) before being lysed using RLT buffer from RNeasy Mini Kit (Qiagen, 74104). RNA was then extracted as per manufacturer’s instructions and RNA concentration was determined using a DA-11 spectrophotometer (DeNovix) at an optical density (OD) ratio of A260/280 nm and stored at -80°C. 1ug of RNA per condition was processed using the protocol listed above, and profiled on an HTA 2.0 (muscle) or Clariom D array (HUVEC, “HTA 3.0”).

## Data Availability

The Global Transcriptomics data as well as the network model data files, code and html data will be released in full after the embargo of publication via ArrayExpress and Zendo for image files and can further be obtained upon reasonable request to the authors

## Contributions

TS, CL, JM, ML, RJB, HR, SP, BEP, WEK, PJA and JAT were responsible for the clinical studies including study design, data collection or data management. CL, KM, CD, PRJM, SP, GS, JPC and JAT were responsible for the Spatial Transcriptomics studies. TCM, JW, HC, CHV, JAT and PJA were responsible for the cell biology studies. JT, TS, IJG and JAS were responsible for the development of informatics solutions for the bulk transcriptomics network analysis models. JT developed the machine learning model for network interpretation with assistance from JG and NMI. MA led the development of the transcriptomic clock model, with assistance from JT and IJG. JT and TS drafted the manuscript, JPC, PJA and WEK refined the draft and the other authors provided further editorial inputs.

## Ethics and Reporting

Full compliance with all normal ethics are reported in detail above within the clinical methods sections. The authors confirm that they have no conflicts of interest, including any payments or services in the past 36 months from a third party that could be perceived to influence, or give the appearance of potentially influencing, the submitted work. Authors confirm that neither we nor our institutions at any time received payment or services from a third party for any aspect of the submitted work.

## Acknowledgements

The studies were funded by European Union Seventh Framework Programme (META-PREDICT, HEALTH-F2-2012-277936); STRRIDE II (NCT00275145) by NHLBI grant HL-057354 and STRRIDE-PD (NCT00962962) by NIDDK DK-081559 and R01DK081559; By the Medical Research Council (G1100015 and MR/Y010329/1); by the NIH, USA (1R56AG061911-01), by NSERC Canada and the Barts Charity, UK (G-002745). We thank Sanjana Sood for her original input into the development of the Custom Chip Definition methodologies as well as the PG students employed to assist with the clinical studies.

## Figures and Figure Legends

**Figure S1.**
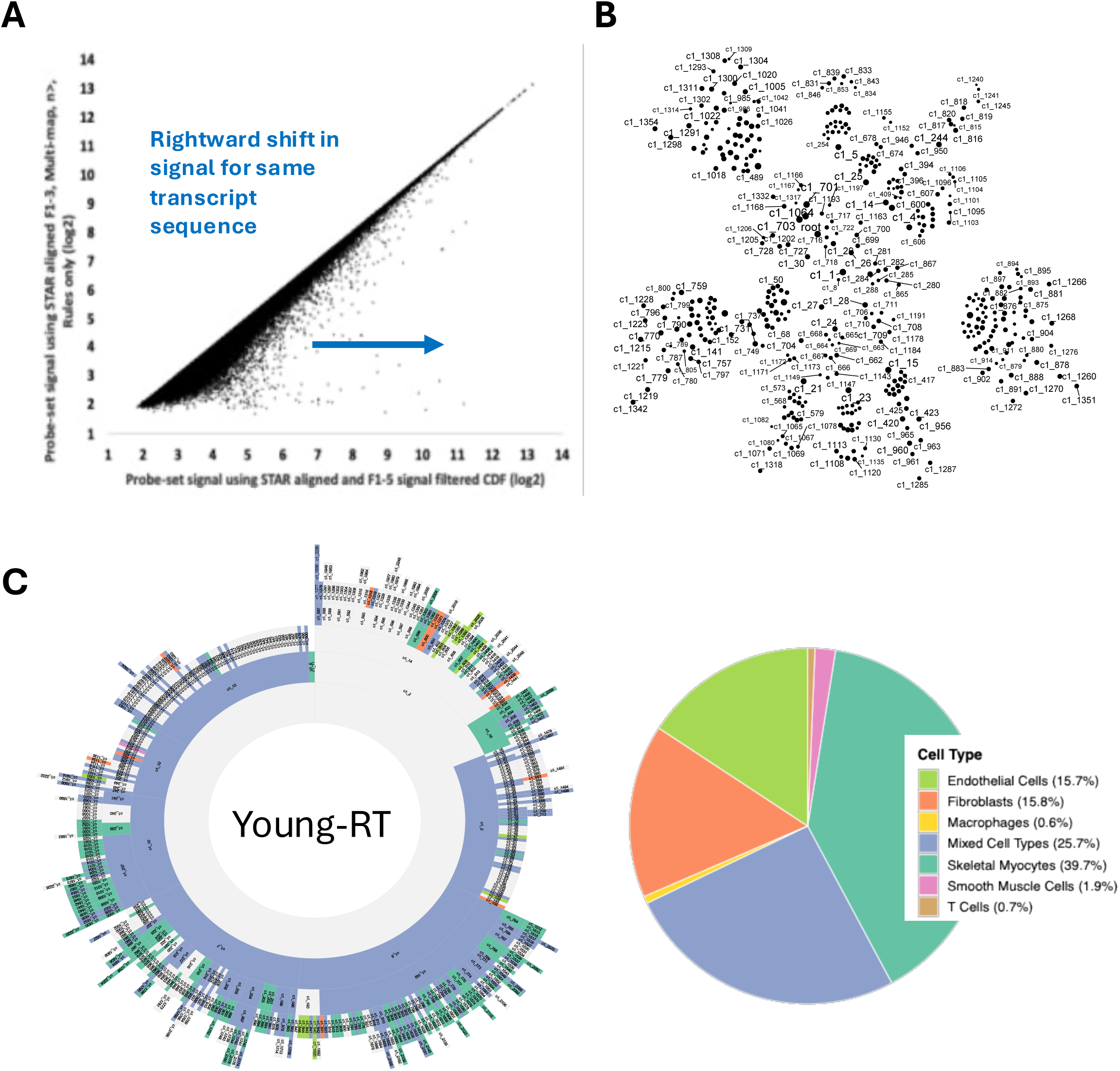
Signal enhancement and old versus new topology format for the human muscle transcriptome. A) Illustration of the optimized data processing pipeline for the exon-based high density arrays that provide more effective coverage of the transcriptome compared with short-read RNAseq. B) Standard MEGENA package network topology plot. C) New sunburst representation of the hierarchal organization of a network constructed from change in gene expression from n=88 people that demonstrated a measurable muscle hypertrophy (from n=144). Each module is colored by its dominant cell type signature with ∼40% of the distal smaller modules being dominated as expected by myocyte gene expression, with the proximal large modules are mixed-cell type profiles.

**Figure S2.**
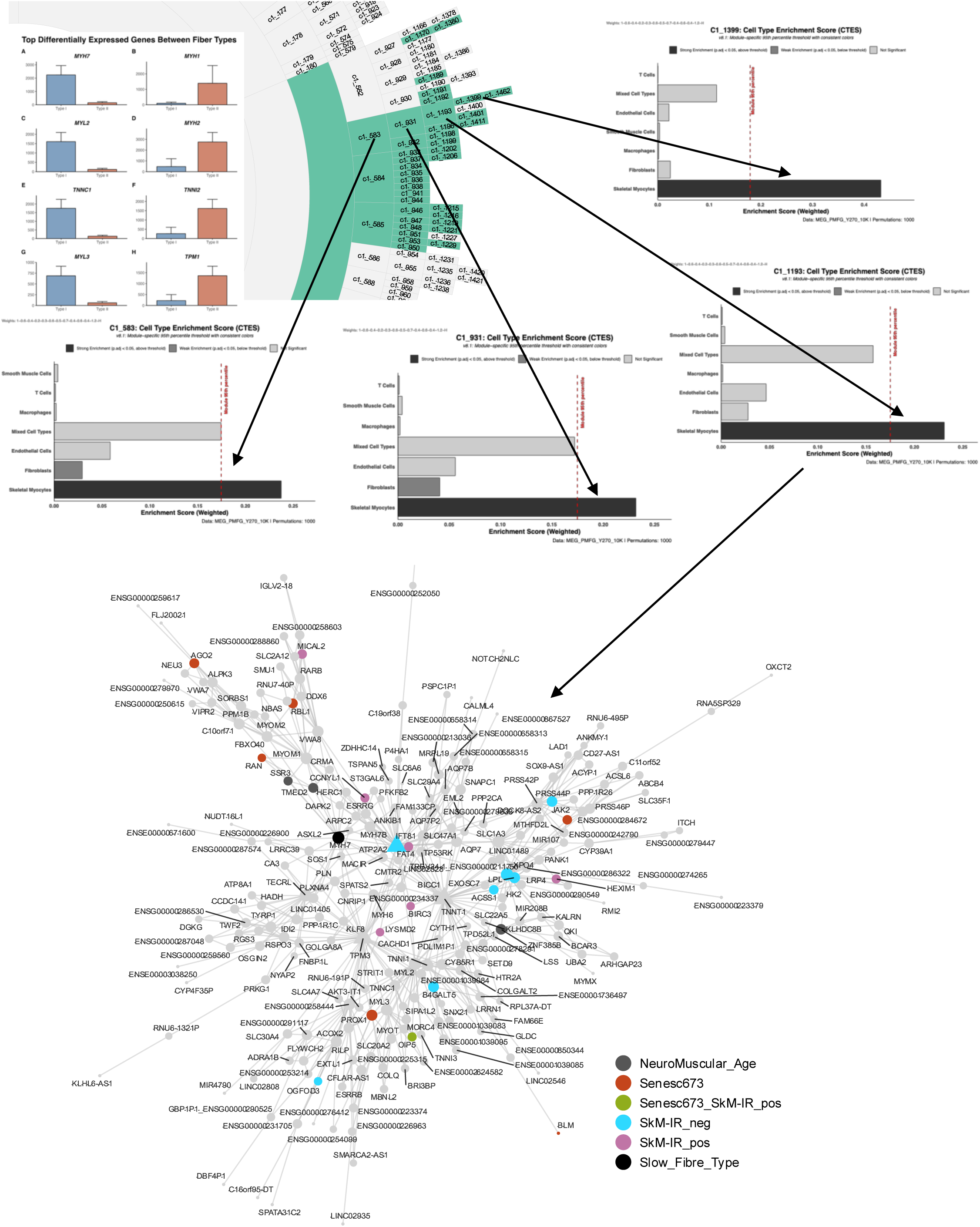
Hierarchal network structure illustrated by the example of Type I muscle fibre modules. Hierarchal network structure illustrated by the example of Type I muscle fibre modules. Note that the module contains several established Type I muscle Fibre marker genes (MYH7, ATP2A2 (hub), TNNT1, TNNI1, TNNC1 and MYH7B). The entire interactive network can be explored using the html version at Zendo (html link)

**Figure S3.**
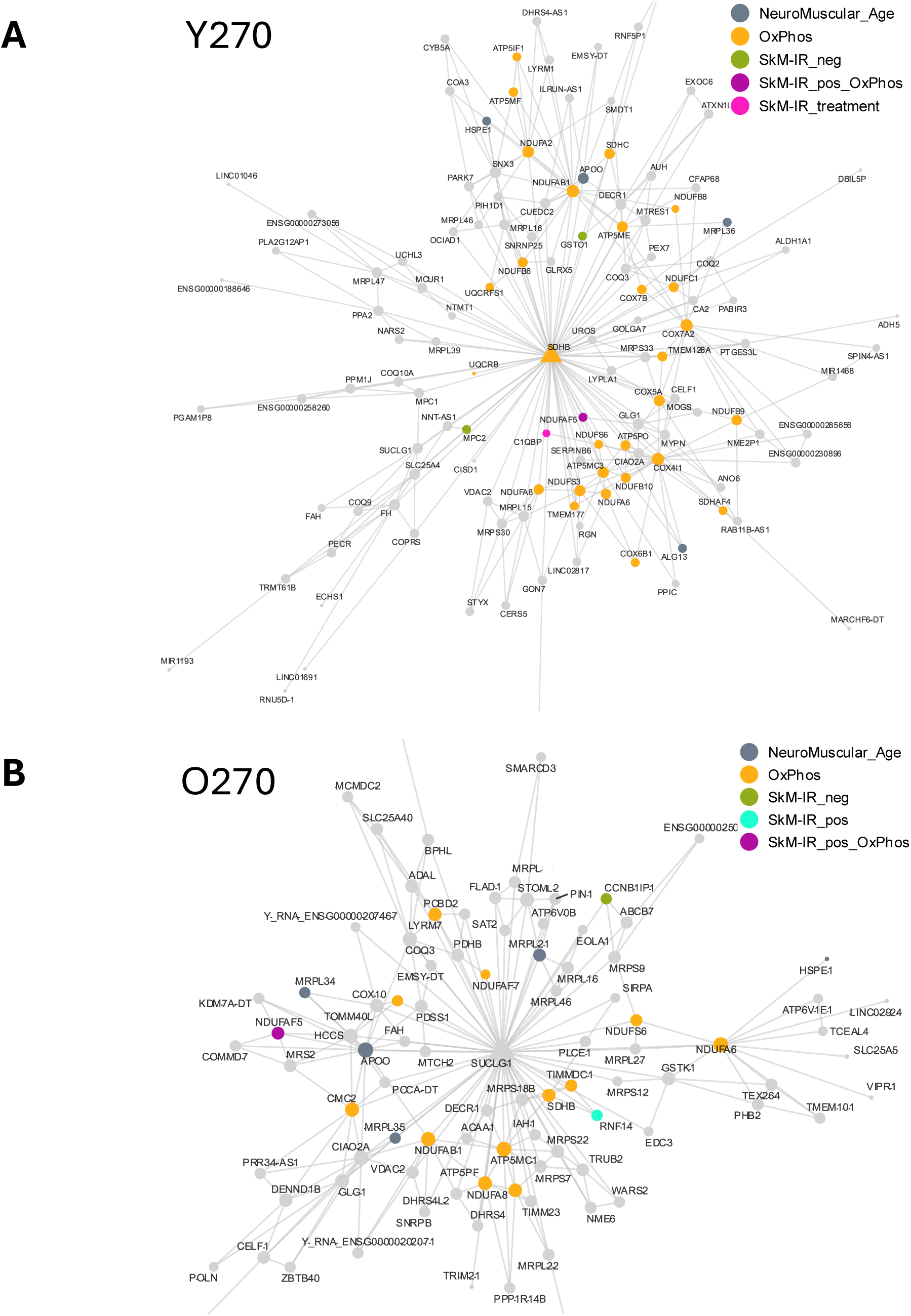
SUCLG1 (succinate-CoA ligase) is a +ve modulator of mitochondrial RNA polymerase, POLRMT.

**Figure S4.**
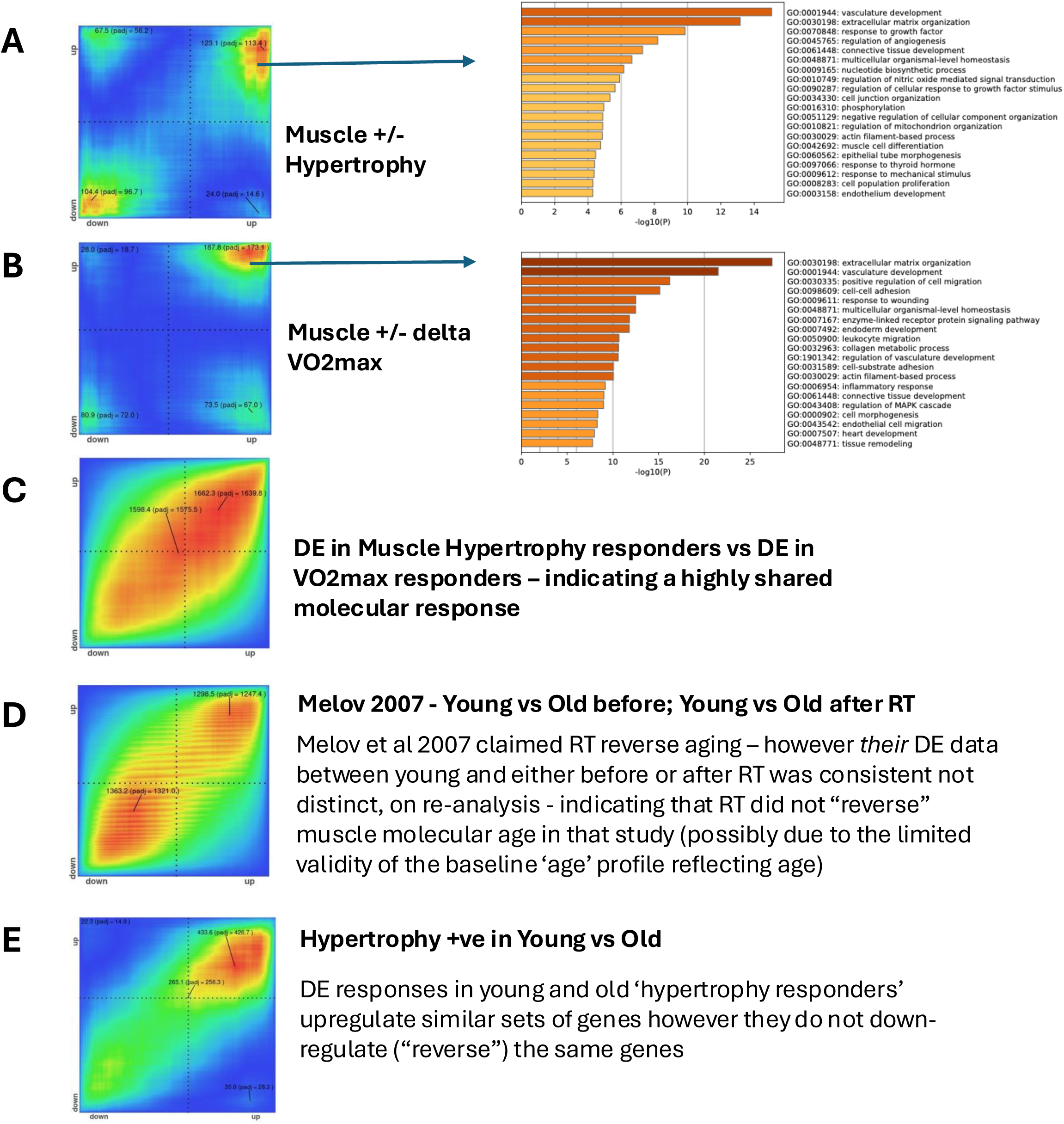
Global transcriptomic contrast of exercise responders and non-responders for hypertrophy or cardiorespiratory capacity. A re-analysis of the Melov 2007 data where reversal of muscle aging was claimed. Contrast of the global DE pattern in young and old responders to hypertrophy

**Fig S5.**
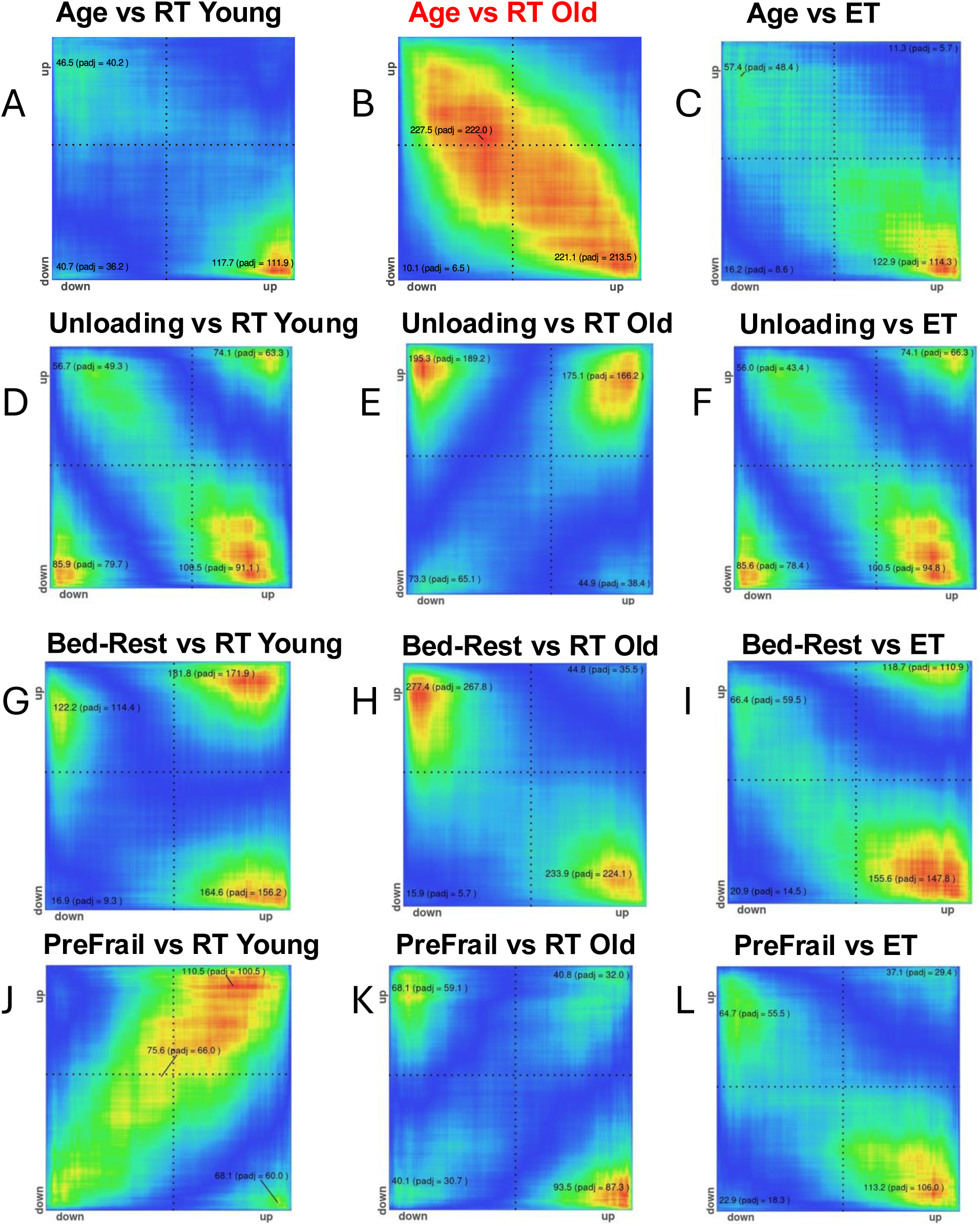
Hypergeometric rank analysis of fold-change values across 28,000 genes. Fold-change values are generated from SMAR analysis and then contrasted using RedRibbon R-Package.

**Figure S6.**
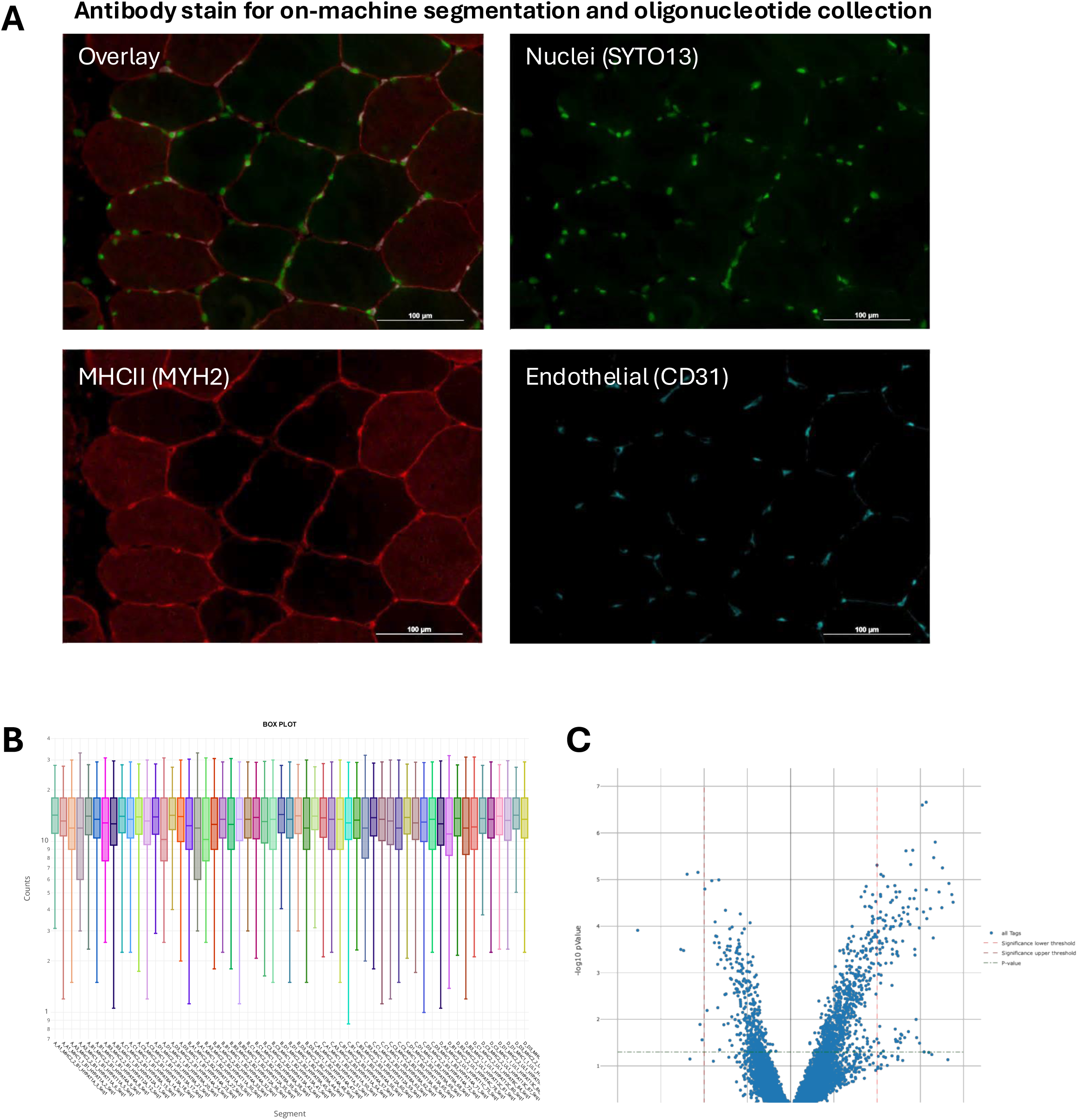
GeoMX Spatial method and QC data. A) segmentation antibody stains. B) 8.6K gene expression after background filter, scale by total AOI area and then quantile normalization. There is no well validated protocol for processing GeoMX data. C) Volcano plot of global differentiation analysis.

**Figure S7.**
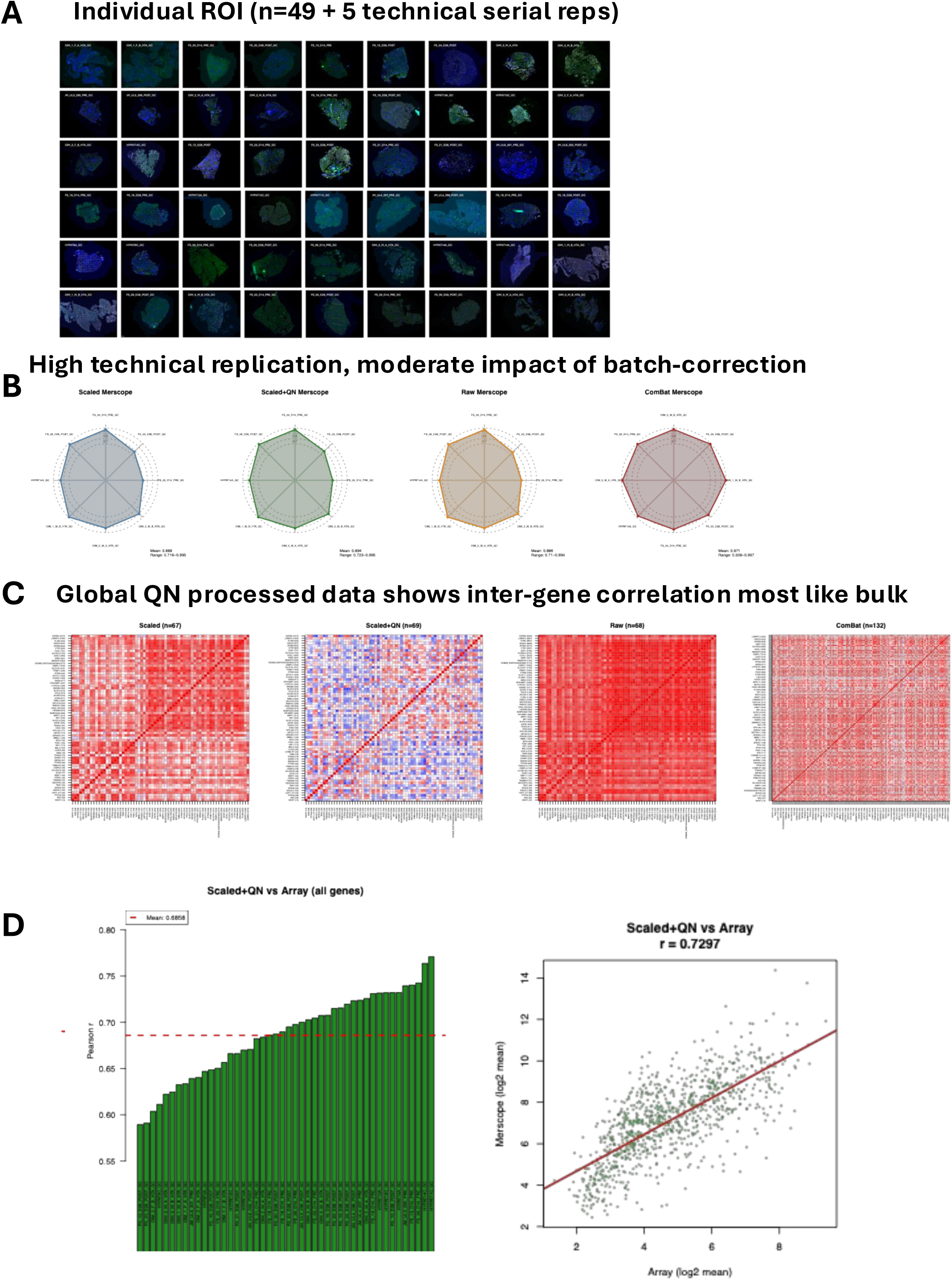
Merscope ROI QC Data

**Figure S8.**
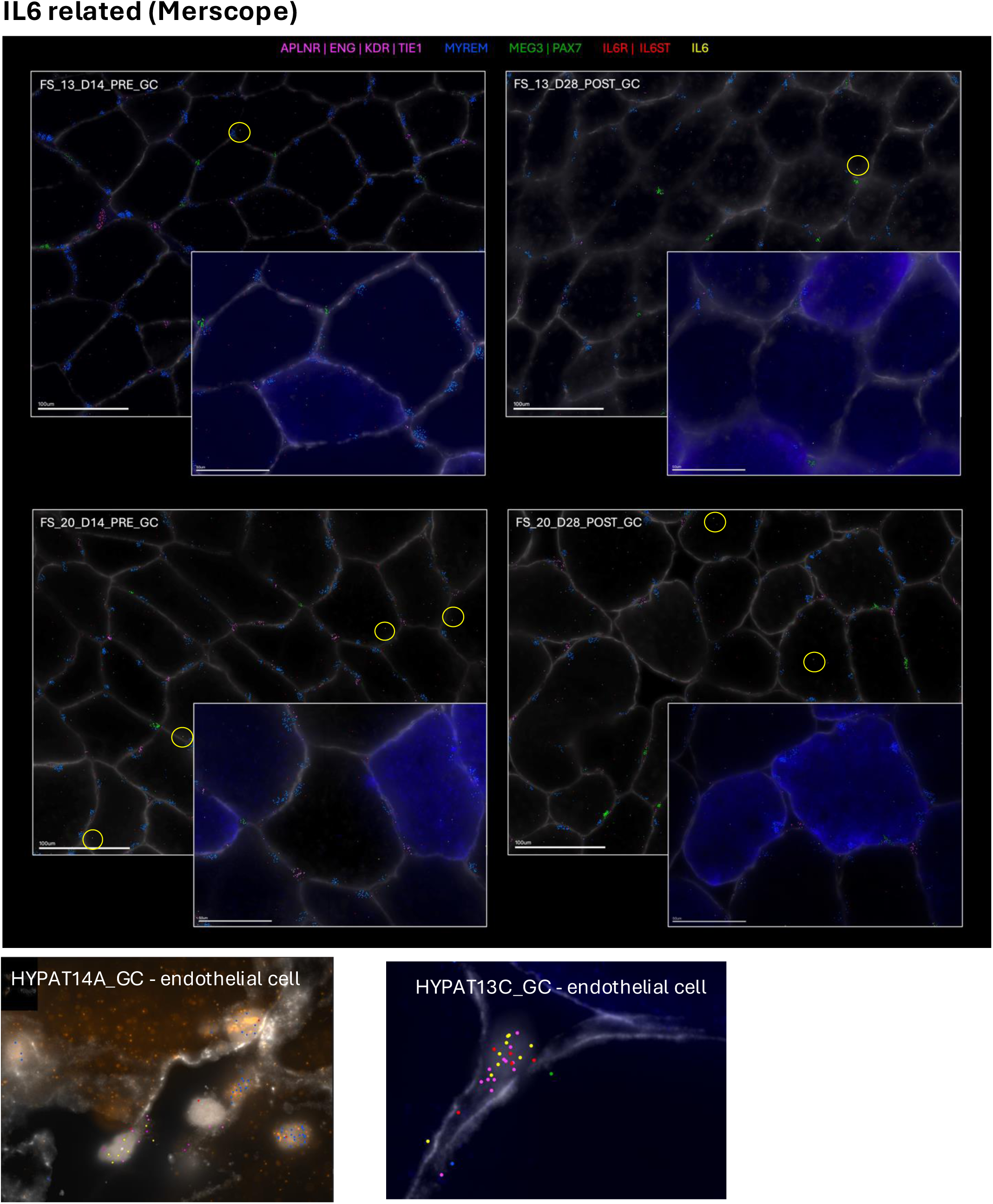
IL6 family. Using Merscope merfish assays we show rare examples of IL6 (yellow) as well as IL6 receptors (red) in four ROIs (with and without muscle use) from samples with the highest total IL6 count out of 54 examples. **A**) Fast (unstained) and slow fibres (MYH7, blue). Mature muscle nuclei (MYREM+; blue) muscle satellite cells (MEG3/PAX7+;green) and endothelial cells (APLNR+/ENG+/KDR+/TIE1+; pink). Overall IL6 is not reliably expressed in human muscle fibres with a single molecule detected in fewer than 1/10^th^ of fibres, a level we cannot distinguish from indirect contamination/background. **B**) In stark contrast, we find *isolated* examples of numerous IL6 mRNA inside endothelial cells in certain biopsy samples (young healthy muscle). These IL6 expressing endothelial cells are rare so they also cannot be the main source of robust IL6 is qPCR experiments using whole biopsy cDNA libraries – which probably reflects immune cells in the biopsy and these are presumably more abundant in post-exercise tissue due to the increased blood perfusion.

**Figure S9.**
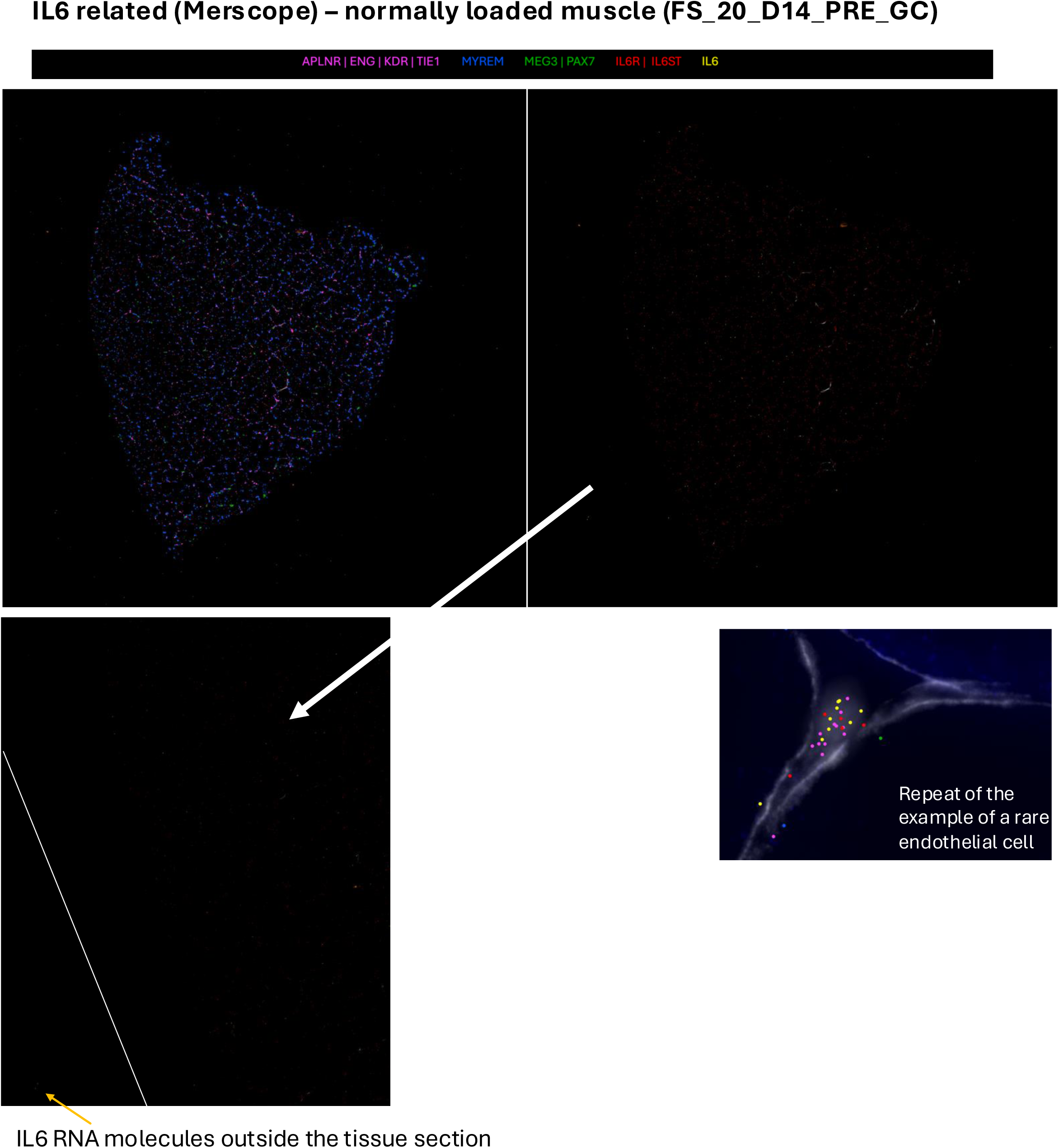
IL6 family. Using Merscope merfish assays we show rare examples of IL6 (yellow) from samples with the highest total IL6 count out of 54 examples. **A**) Low power showing mature muscle nuclei (MYREM+; blue) muscle satellite cells (MEG3/PAX7+;green) and endothelial cells (APLNR+/ENG+/KDR+/TIE1+; pink). IL6 show a single molecule detected in fewer than 1/10^th^ of fibres and can be found outside the tissue section indicative of background or contamination. We show the same example of numerous IL6 mRNA inside endothelial cells in certain biopsy samples (young healthy muscle) for contrast.

**Figure S10.**
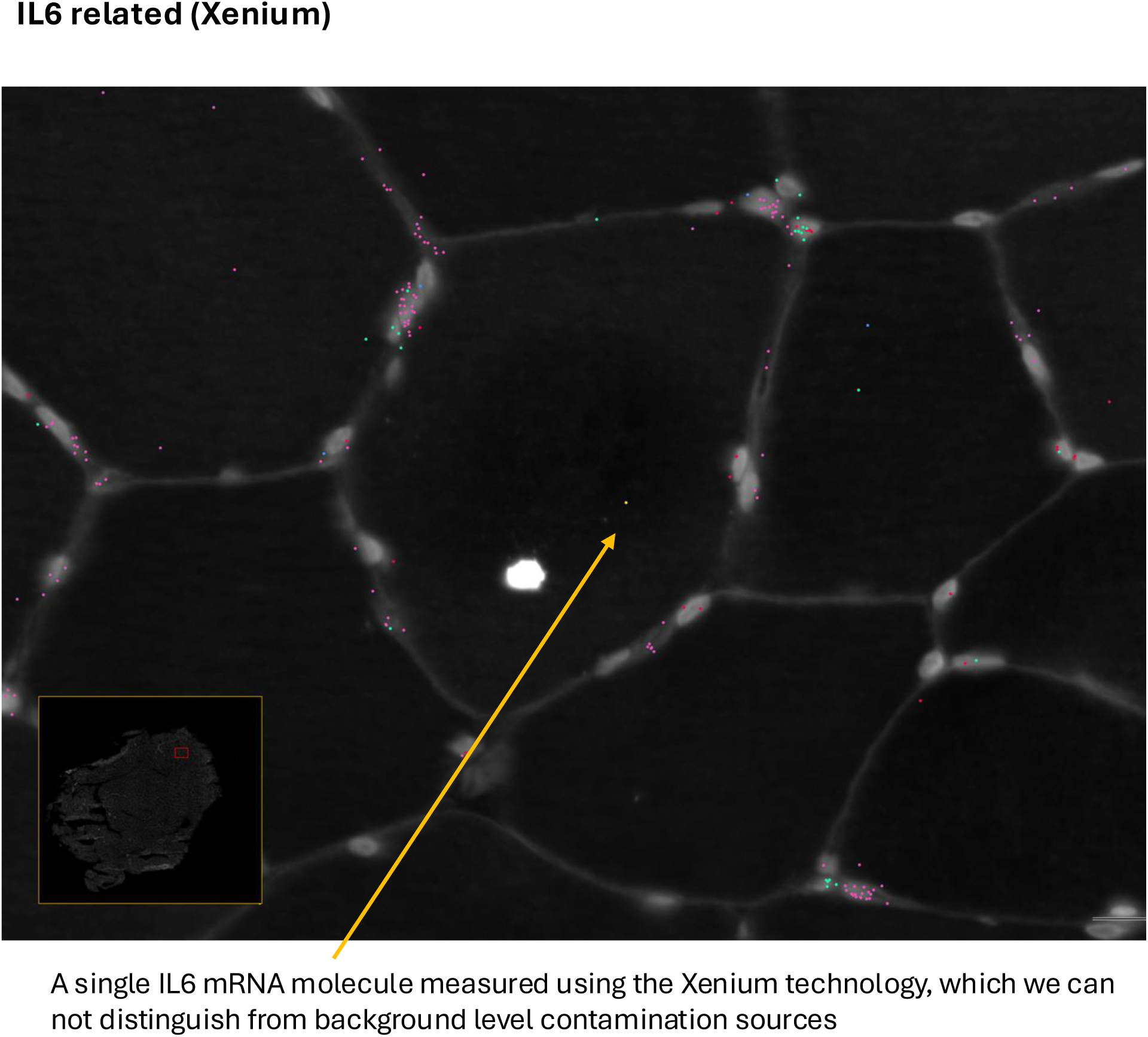
IL6 family. Using the Xenium spatial platform and a 460-plex gene assay (a subset of the Merscope v1 chemistry 960-plex assay) we measure IL6 (yellow) in human muscle. Endothelial cells show specific APLNR, ENG and TIE1 expression (Pink), muscle satellite Cell show MEG3 and PAX7 expression (Green) while mature muscle nuclei expression the noncoding RNA MYREM (Blue). Nuclei are stained with DAPI (show in white as is the cell membrane). Using this second immunofluorescence spatial technology we find no evidence that IL6 is robustly expressed in human muscle cells – with a single IL6 mRNA molecule in one in every ∼20 cells. We cannot rule out these few being non-specific contamination from other cell types. Given the typical signal found using qPCR and cDNA derived from a 20mg tissue biopsy we conclude that IL6 gene expression in muscle tissue is probably derived from immune cells and therefore it is not a myokine.

**Figure S11.**
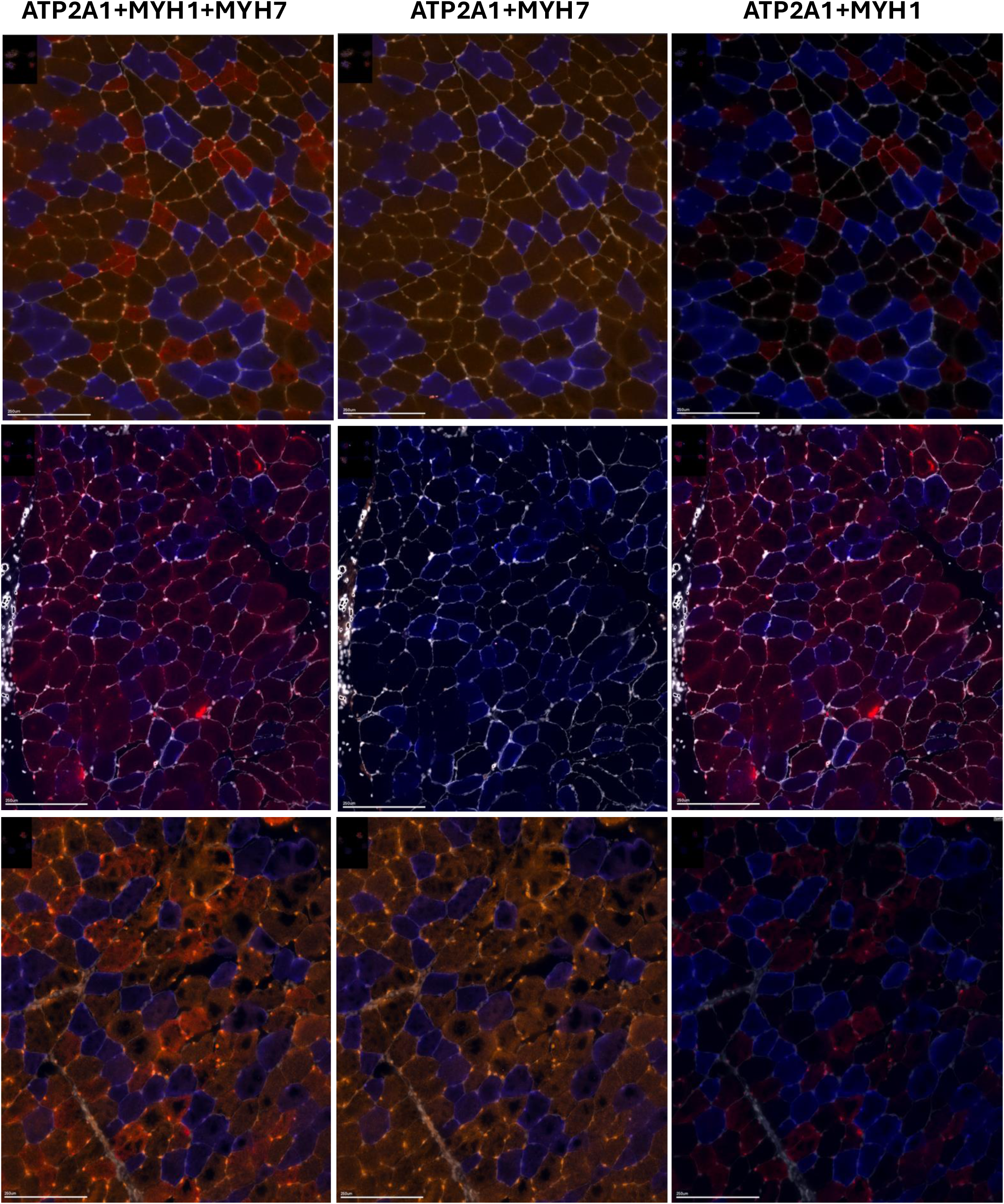
Type I, Type II and IIx staining using the Merscope AUX channels and FISH. Using FISH probes for the three main fibres types, we illustrate the variation in Type II hybrid types we observed using the Merscope technology. This clarifies that Type IIx gene expression is found in human muscle

**Figure S12.**
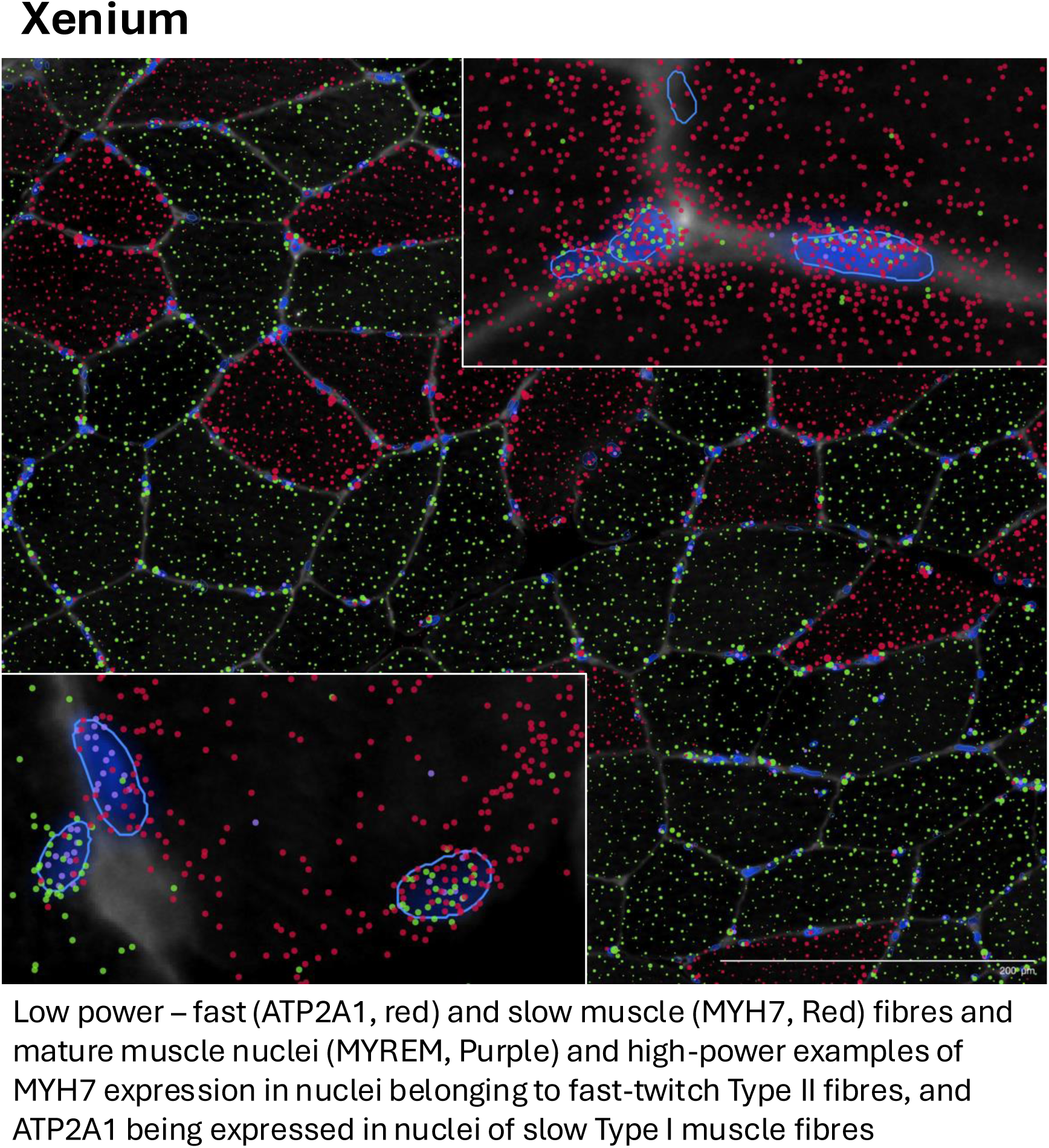
Using the Xenium spatial platform and a 460-plex gene assay (a subset of the Merscope v1 chemistry 960-plex assay) we confirm that some classic cytosolic fibre type specific markers are unsuitable to classify single-nuclei profiles (e.g. MYH7). While MYH7 is, for example, specific to Type I muscle fibres in the cytoplasm, lineage tracing and the present data demonstrate that human muscle nuclei don’t express such genes in a fibre-type specific gene expression pattern and thus can not be utilized to assign snRNAseq profiles to individual fibres types. Only spatial single cell technologies can reliably assign molecular profiles to specific cell types

**Figure S13.**
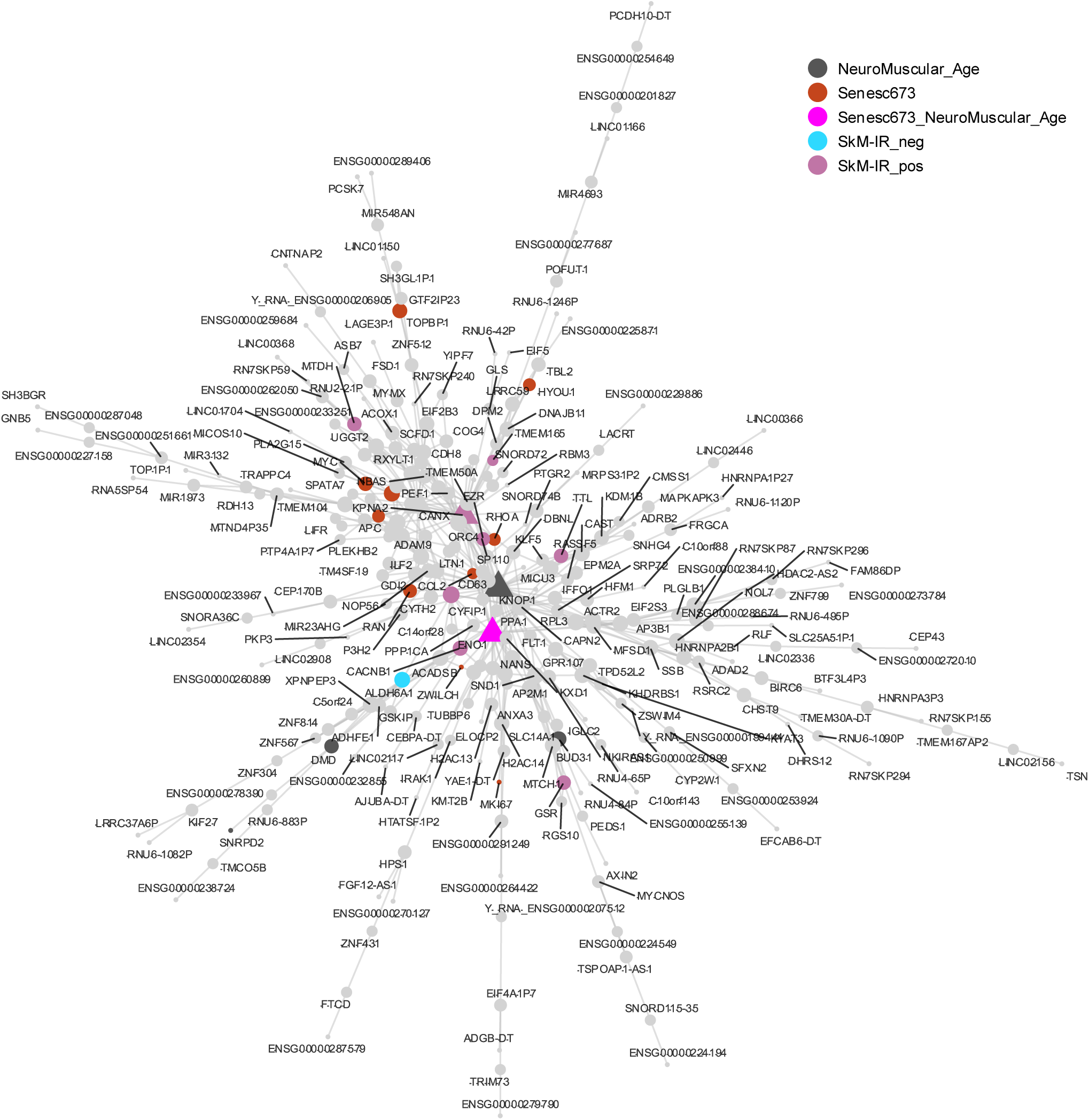

**Figure S14.**
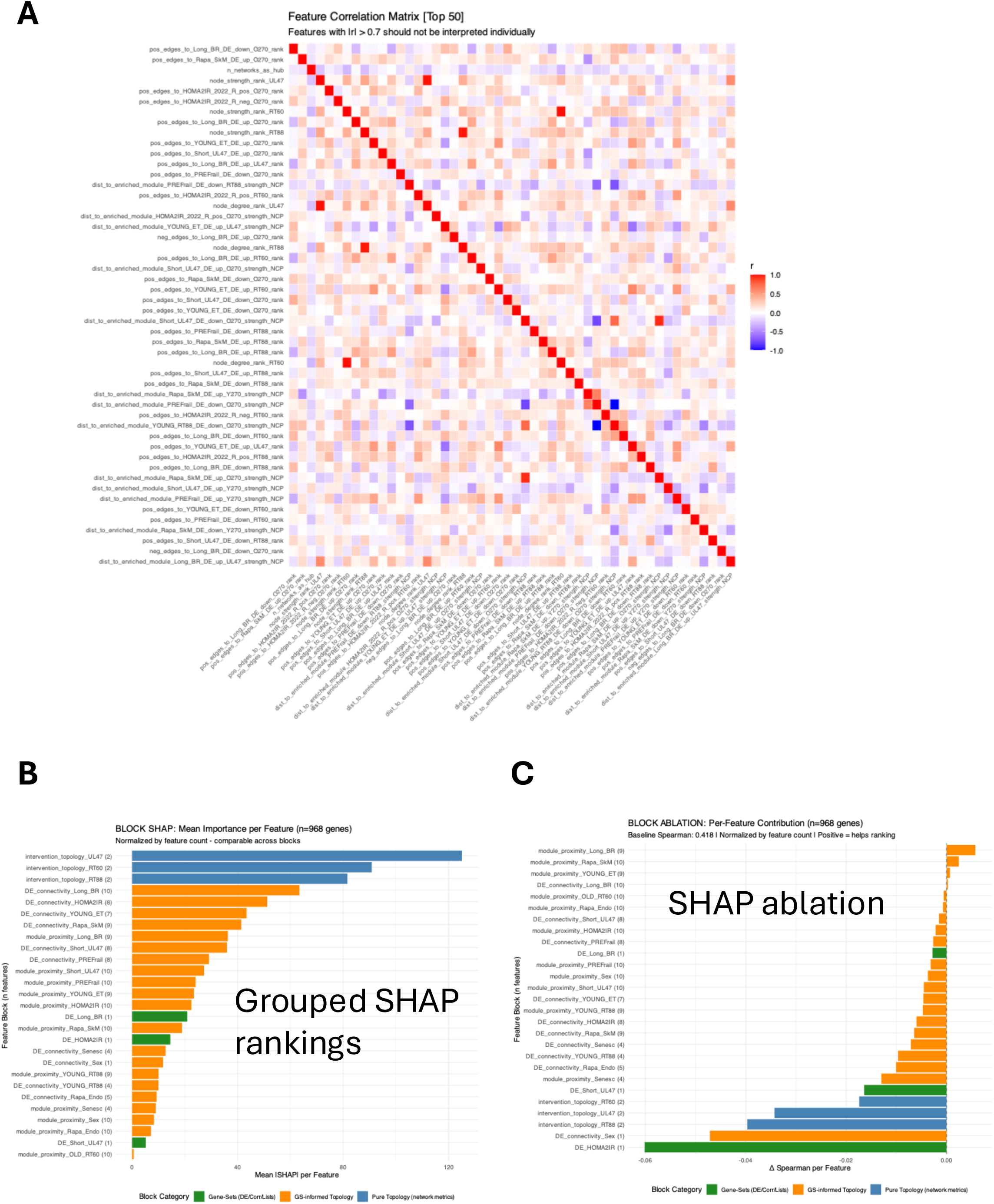

**Supplemental Figure S15.**
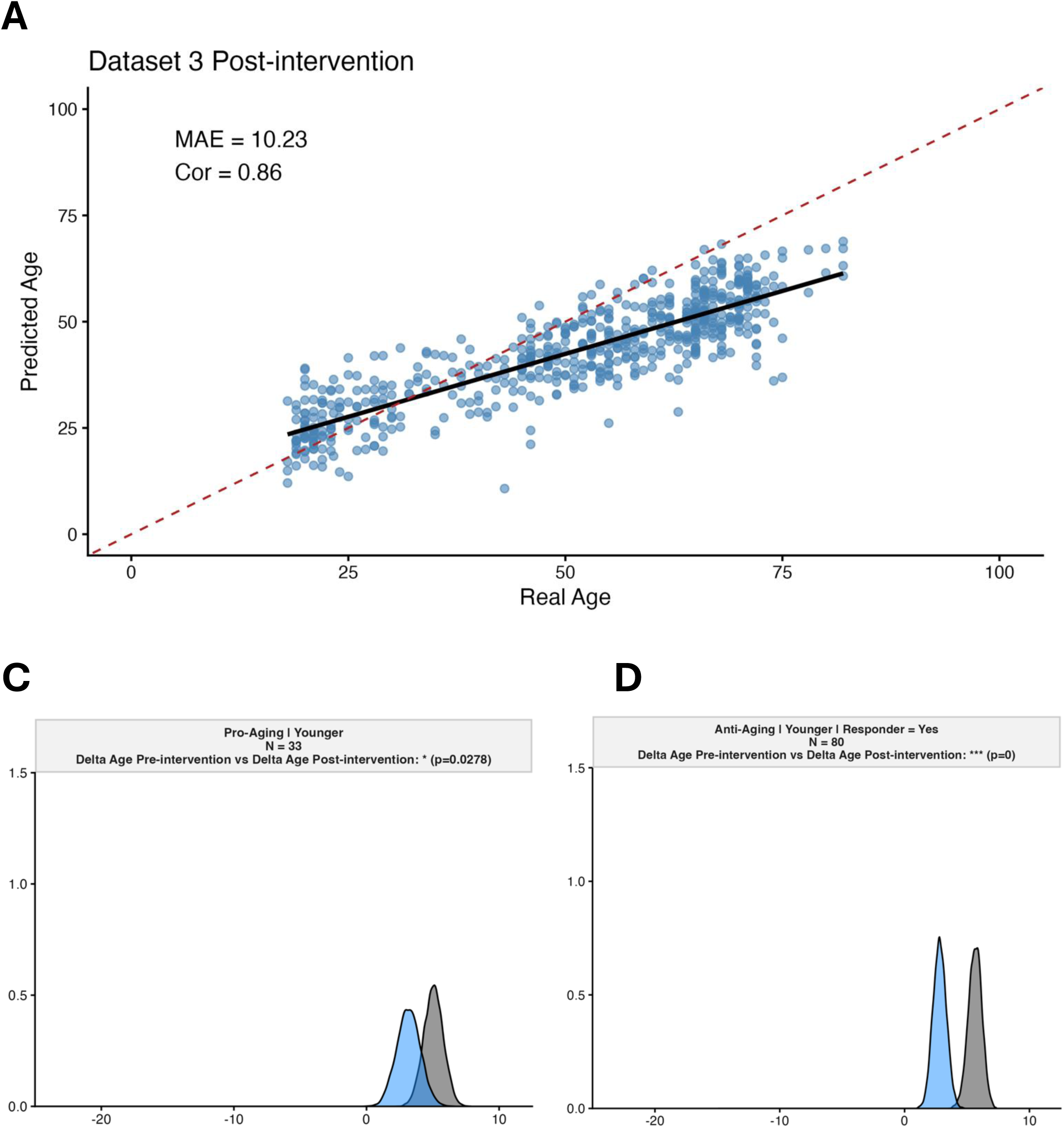
In young people the model performance was more limited, and in fact both pro-aging and anti-aging interventions resulted in a shift in the predicted age distribution. Given the hypothesis and background epidemiology, these analysis, from smaller data sets lack biological plausibility.

